# A high-throughput microfluidic nano-immunoassay for detecting anti-SARS-CoV-2 antibodies in serum or ultra-low volume dried blood samples

**DOI:** 10.1101/2020.10.07.20208280

**Authors:** Zoe Swank, Grégoire Michielin, Hon Ming Yip, Patrick Cohen, Diego O. Andrey, Nicolas Vuilleumier, Laurent Kaiser, Isabella Eckerle, Benjamin Meyer, Sebastian J. Maerkl

## Abstract

Novel technologies are needed to facilitate large-scale detection and quantification of severe acute respiratory syndrome coronavirus 2 (SARS-CoV-2) specific antibodies in human blood samples. Such technologies are essential to support seroprevalence studies, vaccine clinical trials, and to monitor quality and duration of immunity. We developed a microfluidic nano-immunnoassay for the detection of anti-SARS-CoV-2 IgG antibodies in 1024 samples per device. The method achieved a specificity of 100% and a sensitivity of 98% based on the analysis of 289 human serum samples. To eliminate the need for venipuncture, we developed low-cost, ultra-low volume whole blood sampling methods based on two commercial devices and repurposed a blood glucose test strip. The glucose test strip permits the collection, shipment, and analysis of 0.6 *µ*L whole blood easily obtainable from a simple fingerprick. The nano-immunoassay platform achieves high-throughput, high sensitivity and specificity, negligible reagent consumption, and a decentralized and simple approach to blood sample collection. We expect this technology to be immediately applicable to current and future SARS-CoV-2 related serological studies and to protein biomarker diagnostics in general.

## Introduction

The emergence of a new coronavirus at the end of 2019, termed severe acute respiratory syndrome coro-navirus 2 (SARS-CoV-2), led to an unprecedented global public health crisis [1]. Over a year later, it is estimated that SARS-CoV-2 infected over 100 million people worldwide and two million people died of coronavirus disease 2019 (COVID-19) caused by SARS-CoV-2 [2]. As SARS-CoV-2 causes mainly mild disease or infection presents without symptoms, many cases are not captured by direct testing in the acute phase of disease [3]. However, to estimate infection fatality rate and guide public health decisions, it is of utmost importance to establish the true spread or prevalence of the virus by identifying how many people have been exposed [4, 5].

Detection of anti-SARS-CoV-2 antibodies using highly sensitive and specific assays can help answer these questions. Several seroprevalence studies have already been conducted, demonstrating rather low seroprevalence rates even in areas that were severely affected [6, 7, 8, 9]. Such data show that herd immunity through natural infection is far from being reached. However, these studies are merely snapshots of an evolving situation, both in time and space. Therefore, there is a sustained need for seroprevalence studies to be continuously conducted in order to monitor virus spread and to keep policy makers informed. In addition, tens to hundreds of thousands of blood samples will need to be tested to determine antibody titers for each SARS-CoV-2 vaccine phase 3 clinical trial and a large number of samples will need to be tested to monitor immune responses after a vaccine has been approved and rolled-out. Such studies are cumbersome and expensive to perform as they require large-numbers of serum samples to be obtained by venipuncture and analysed by highly sensitive and specific immunological assays to classify samples or to provide quantitative information on antibody titers.

Several assays, such as enzyme-linked immunosorbent assays (ELISA) or chemiluminescent immunoassays (CLIA), are commercially available, but mainly rely on serum drawn by venipuncture. These tests are also rather expensive, with reagent costs on the order of 3-10 USD per test. Alternatively, in-house ELISAs are difficult to standardize and require high amounts of recombinant antigen, usually around 100ng per sample [5]. Other recently developed methods such as miniaturized high-throughput ELISAs that use low microliter volumes suffer from lower sensitivity [10], and ultra-sensitive assays based on digital ELISA have a low sample throughput of 68 tests per hour [11]. The comparatively high cost of these assays and the reliance on serum samples taken by venipuncture are considerable hurdles to performing large-scale studies under normal circumstances, but especially so during a pandemic, when sample collection can put clinical staff and study participants at risk. Lateral flow assays (LFAs) can be performed at the point of care or at home requiring only a “drop” of whole blood, but the sensitivity and specificity of these assays is often insufficient [12, 13, 14] and LFAs are relatively expensive at ∼22 USD per test. Furthermore, LFAs provide test results but no blood samples are being collected which could be used for follow-up analyses.

There is therefore a clear need for new technologies to supersede existing methods such as ELISA, CLIA, and LFAs. Novel technologies should be capable of high-throughput, low-reagent consumption, low-cost per test, high sensitivity and specificity, and be compatible with ultra-low volume whole-blood samples in the low or even sub-microliter range that can be obtained via a simple fingerprick. Biomarker detection using dried whole blood on filter paper or other devices would have tremendous advantages as it can be collected by untrained individuals at home. The samples could then be conveniently shipped by regular mail at ambient temperature to a central laboratory for analysis and test results returned electronically via a mobile app or email.

In this study we developed and validated a nano-immunoassay (NIA) that analyses 1024 samples in parallel on a single microfluidic device the size of a USB stick. NIA reagent consumption and corresponding costs are roughly 1000 times less than a standard ELISA. Based on the analysis of 134 negative and 155 positive control sera, NIA achieved a specificity of 100% and a sensitivity of 98% when RT-PCR was used as a reference. NIA performed as well as a standard ELISA for samples obtained more than 20 days post onset of symptoms, and performed better than ELISA for samples obtained less than 20 days past onset of symptoms, indicating that NIA is more sensitive than ELISA for the analysis of early timepoint samples with lower antibody titers. We go on to demonstrate that NIA can be used to measure anti-SARS-CoV-2 antibodies in ultra-low volume dried whole blood samples, eliminating the need for venipuncture blood collection. We tested two commercial blood collection devices: Neoteryx’s Mitra® and DBS System SA’s HemaXis™ DB10, and show that it is possible to repurpose low-cost and widely available blood glucose test strips for sample collection and shipment. Samples could be stored up to 6 days at room temperature with minimal sample degradation and all three methods yielded better results than an ELISA performed on standard serum samples collected from the same individuals.

## Results

### Nano-immunoassay development

We adapted a MITOMI based [15, 16] 1024 NIA device previously applied to vaccine adjuvant screening [17] and the detection of inflammatory and cancer related protein biomarkers in serum [18] to the detection of anti-SARS-CoV-2 IgG antibodies. The NIA device described here is a simplified version of the original 1024 serum analyzer chip and more similar to the original MITOMI device with slightly enlarged chambers to accommodate spotted patient samples. The microfluidic device is a standard two-layer polydimethylsiloxane (PDMS) device [19], consisting of a flow and a control layer. Fluids in the flow layer can be manipulated with pneumatic valves formed by the control layer. The device contains 1024 unit cells, each consisting of an assay and a spotting chamber (Figure 1A). Patient samples are spotted onto an epoxy-coated glass slide using a contact-printing microarray robot, on top of which the PDMS device is aligned and bonded. Patient samples are initially isolated by actuating the neck valve and the surface of the assay chamber is patterned with biotinylated bovine serum albumin (BSA-biotin) and neutrAvidin, leaving a circular surface region coated with neutrAvidin beneath the MITOMI button. Afterwards, biotinylated anti-His antibody is flowed through the device, enabling the subsequent immobilization of a His-tagged SARS-CoV-2 antigen. Patient samples are then solubilized and the sandwich valves are closed, allowing any SARS-CoV-2 specific antibodies to diffuse into the assay chamber and bind the surface immobilized antigen. The MITOMI button is closed following an incubation period and any unbound material is washed away. A secondary antibody labeled with phycoerythrin (PE) is flowed to detect the presence of antibodies bound to the SARS-CoV-2 antigen (Fig. 1B, C).

**Figure 1:**
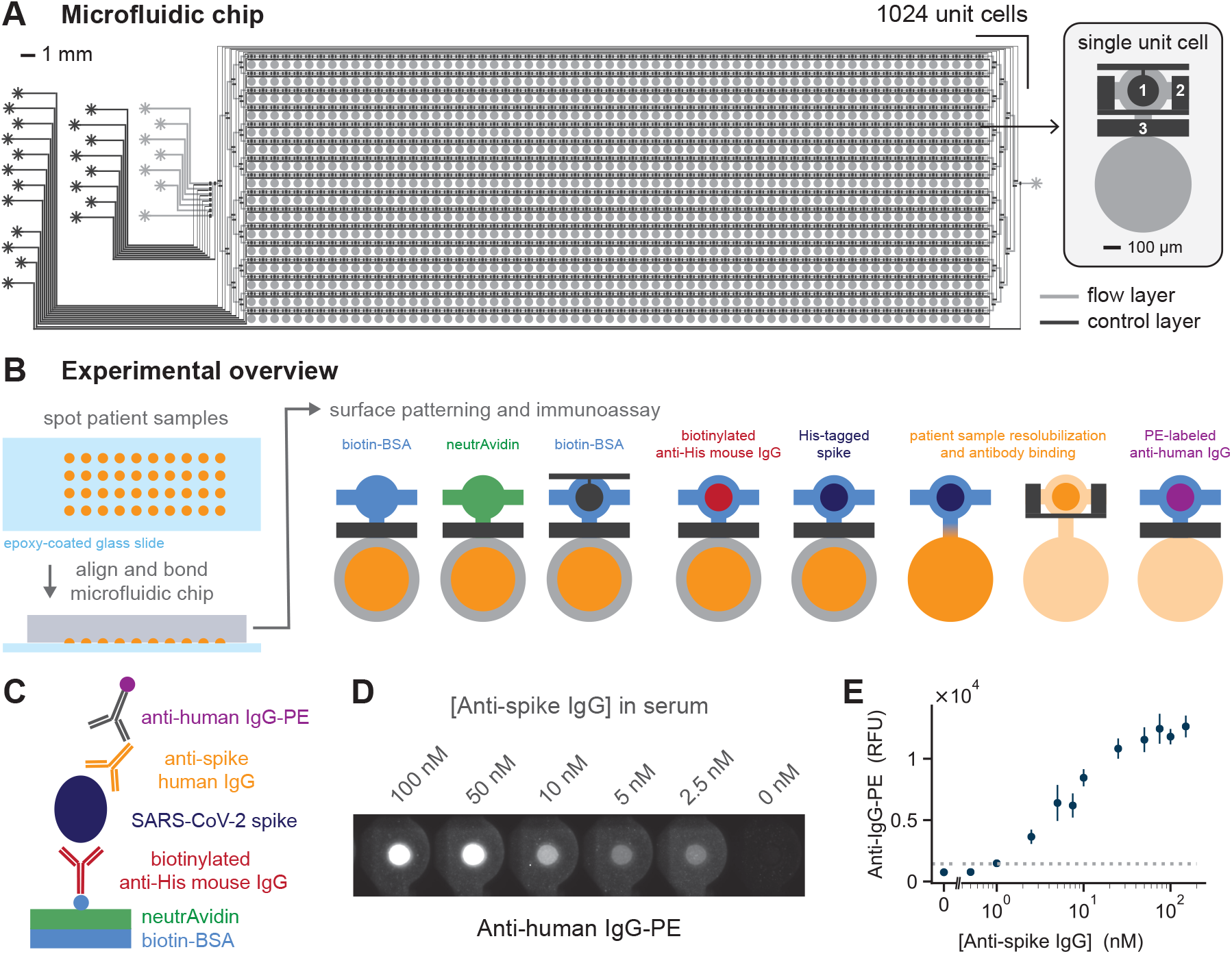
High-throughput microfluidic nano-immunoassay for anti-SARS-CoV-2 antibody detection. **(A)** The two-layer microfluidic chip design consists of 1024 unit cells. Each unit cell in turn contains two sections: an immunoassay chamber (top) and a spotting chamber (bottom). Control valves include the button (1), sandwich (2) and neck (3) valves. **(B)** A schematic of the experimental process, starting with the spotting of patient samples, followed by chip alignment and bonding, biotin-BSA and neutrAvidin surface patterning, and lastly the immunoassay for detection of anti-SARS-CoV2 antibodies. Surface patterning and immunoassay are shown for a single unit. During the experiment all unit cells are processed in parallel. **(C)** Schematic of the on-chip sandwich immunoassay. **(D)** Fluorescence images of anti-human IgG-PE signal for a given concentration of anti-spike antibodies present in human serum. **(E)** Quantification of the anti-human IgG-PE signal for a range of anti-spike concentrations spotted. The dashed horizontal line indicates the LOD.

We optimized the NIA for detection of anti-spike IgG antibodies in human serum by first testing several different spike antigen variants and identified the in-house produced trimerized full-length spike protein as the most suitable antigen (Fig. S1). The use of trimerized full length spike and spike-receptor binding domain (RBD) both resulted in high signal intensities and low-background. We decided on using the trimerized full-length spike protein as it most closely resembles the natural conformation of the SARS-CoV-2 spike protein. As a proof-of-concept we added chimeric anti-spike antibody at different concentrations into human serum, spotted each dilution, and showed that NIA could quantify anti-spike antibody concentrations (Fig. 1D, E). Based on this sandwich immunoassay we estimate that our limit of detection (LOD) is around 1 nM IgG. To permit the analysis of patient sera in a standard, biosafety level 1 (BSL-1) laboratory we developed a simple treatment protocol that renders the sera BSL-1 compatible. This was achieved by conducting a short heat-inactivation step, followed by the addition of Triton X-100 to a final concentration of 1% [20]. Dried whole blood samples could be safely handled in a BSL-1 environment without requiring any pre-treatment steps.

### Nano-immunoassay validation

We validated the high-throughput NIA for the detection of SARS-CoV-2 anti-spike IgG antibodies with 289 serum samples, collected from 155 PCR-confirmed SARS-CoV-2 infected individuals and 134 prepandemic negative samples collected in 2013/14 and 2018. Different serum sample dilutions, ranging from no dilution to a 1:256 dilution, were spotted to determine the optimal value (Fig. S2). NIA achieved a maximum specificity and sensitivity of 100% and 98%, respectively, at a serum dilution of 1:8 (Fig. 2A, S2) and a receiver operator characteristic (ROC) curve with an area under the curve (AUC) equal to 0.99 (Fig. 2B). In parallel, ELISA was used to validate the presence of anti-spike(S1) IgG in the same patient samples (Fig. 2C) resulting in a good correlation (R^2^=0.87) between NIA and ELISA measurements (Fig. 2D). As the serum samples were obtained from patients at different time points post onset of symptoms we plotted the NIA results according to when samples were obtained (Fig. 2E). We also analyzed NIA and ELISA results split into samples obtained on or before 20 days post onset of symptoms (DPOS) and samples obtained after 20 days post onset when antibody responses are usually fully developed (Fig. 2A, C). If DPOS was not known for a given patient, then days post diagnosis (DPD) was used instead. Performance in terms of specificity and sensitivity between NIA and ELISA are similar for samples obtained after 20 days past onset of symptoms, but NIA outperforms ELISA for samples collected on or before 20 days post onset in terms of sensitivity by 13 percentage points (Fig. 2F). In addition, four other commercially available assays were used to detect anti-SARS-CoV-2 IgG antibodies present in a subset of the COVID-19 patient serum samples. For samples collected after 20 days post onset NIA performed as well as 2 other commercial assays and outperformed 2 of these assays in terms of sensitivity. Furthermore, NIA outperformed all 5 commercially available assays in terms of sensitivity for samples collected less than 20 days post onset by at least 9 percentage points or more (Fig. S3).

**Figure 2:**
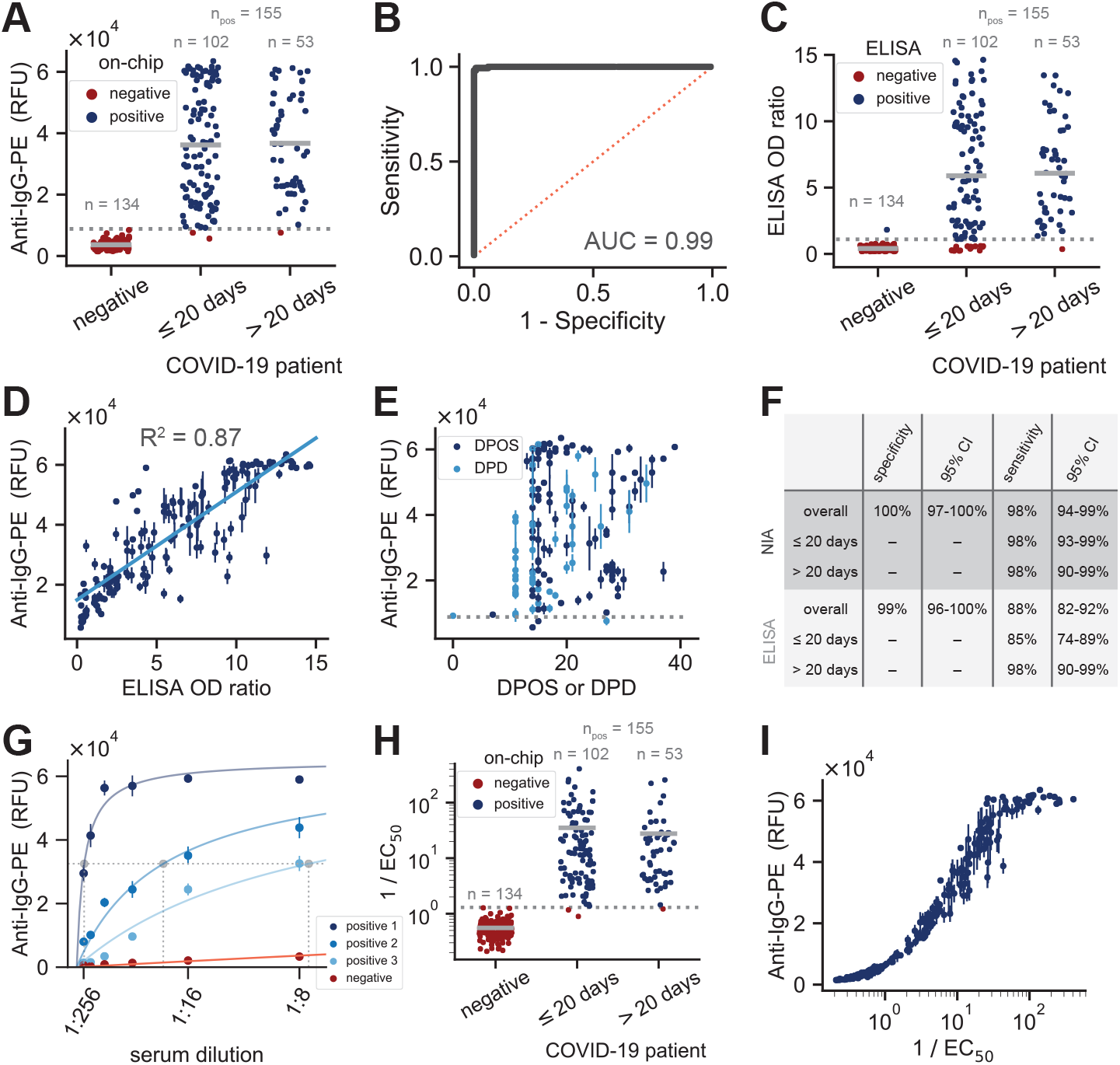
Nano-immunoassay validation. **(A)** NIA anti-IgG-PE signal for serum samples obtained from SARS-CoV-2 negative individuals (obtained in 2013/14 and 2018) or SARS-CoV-2 RT-PCR confirmed positive patients. Measurements shown are for a 1:8 dilution of each serum sample. Data points represent mean values of 3 replicates from a single chip. A total of 6 devices were run to collect this data. **(B)** ROC curve corresponding to the measurements shown in panel A. **(C)** ELISA OD ratios for the same serum samples as in A. **(D)** Correlation between NIA and ELISA measurements. Data points represent means ± SD (*n* = 3). **(E)** NIA measurements plotted versus days past onset of symptoms or days post diagnosis. Data points represent means ± SD (*n* = 3). **(F)** Specificity and sensitivity for NIA and ELISA. The dashed grey lines in A, C, and E indicate the cutoff used for the specificity and sensitivity calculations. **(G)** Examples of curve fits for three positive and one negative patient sample. The horizontal dashed line indicates half maximal binding and the three dashed vertical lines correspond to the *EC*_50_ values determined for each of the positive samples. **(H)** 1*/EC*_50_ categorized according to whether the sample was negative or from an RT-PCR confirmed positive patient. **(I)** Anti-IgG-PE signal for a 1:8 dilution of each serum sample versus 1*/EC*_50_.

Each sample dilution was tested in triplicate, enabling us to test up to 336 different samples or sample dilutions on a single device. In order to determine if it would be possible to further increase throughput by reducing the number of replicates we randomly selected one or two of the three 1:8 dilution replicates and calculated ROC curves, specificity, and sensitivity (Fig. S4). We found that using duplicates only slightly reduced sensitivity from 98% to 97%, whereas using a single measurement lowered sensitivity to 95%, all the while retaining a 100% specificity. Given that using duplicate measurements led to only a minimal change in sensitivity, it will be possible to increase throughput to 512 samples analyzed in duplicate per device. Additional gains in throughput are theoretically possible by scaling the device itself, with MITOMI devices containing up to 4160 unit cells having been demonstrated in the past [21].

To assess device to device reproducibility we tested all serum samples (1:8 dilution) on two separate devices which resulted in a correlation of *R*^2^ = 0.98 (Fig. S5A). We then correlated the measurements from one of those devices to the measurements made on six other devices and again observed a good correlation of *R*^2^ = 0.95 (Fig. S5B). In the first case the same sample solution was used for spotting two devices, whereas in the second case the two measurements being compared came from separately prepared patient sample dilutions. Additionally we compared the anti-IgG-PE signal measured for a reference serum dilution series on three separate devices and computed an average coefficient of variation equal to 11.7% for the dilutions measured (Fig. S5C, D).

To determine quantitative antibody titers in each serum sample, we fit data from the full dilution series to a saturation binding curve model, enabling us to extract the dilution equivalent to the half-maximum signal for each serum sample (*EC*_50_) (Fig. 2G, S6). Using the calculated 1*/EC*_50_ value, we again analyzed the NIA results, resulting in a specificity and sensitivity of 100% and 98%, respectively (2H). Lastly, we compared the 1*/EC*_50_ values to values obtained from a single 1:8 dilution and found an excellent correspondence between those two measurements (Fig. 2I), indicating that a single measurement returns accurate quantitative information on antibody concentrations. Only at very high antibody titers does the signal saturate for the 1:8 dilution. Using two measurements of a 1:8 and one slightly higher dilution could therefore maximize throughput, specificity, and sensitivity of NIA, and return accurate antibody titer values.

### Ultra-low volume whole blood collection and analysis

The NIA platform proved to be highly specific and sensitive when applied to standard serum samples. We recognized that use of serum samples limits the applicability of our and other methods to samples collected by venipuncture, which has to be performed by trained personnel in hospitals or point of care settings. Venipuncture requires large volumes of blood in the 5 mL range, considerable downstream sample processing, and a cold chain from point of sample collection to point of analysis. Large-scale seroprevalence studies are thus challenging to conduct, especially when including remote or large geographic areas. Establishing a large-scale, low-cost, and widely accessible serology platform necessitates the development of venipuncture alternatives. For this reason, we developed a sample collection and processing pipeline that enables us to carry out NIA with ultra-low volume, dried whole blood samples obtainable by a simple fingerprick. We tested three different methods to collect, ship, process, extract, and analyze dried whole blood samples (Fig. 3). We tested two commercially available devices that collect 10 *µ*L of whole blood: Neoteryx Mitra® and DBS System SA HemaXis™ DB10. We also explored the possibility of repurposing existing blood glucose test strips (Medisana, MediTouch 2) for whole blood collection and shipping. The glucose test strips collect 0.6 *µ*L of whole blood which is approximately twenty times less than what is required by the commercial devices and 10,000 times less blood than obtained by venipuncture. Furthermore, blood glucose test strips are cheap at less than 0.5 USD per strip and widely available, potentially avoiding any supply bottlenecks during a pandemic. The two commercial devices by comparison cost 5-10 USD or 1.25 - 2.5 USD per sample.

**Figure 3:**
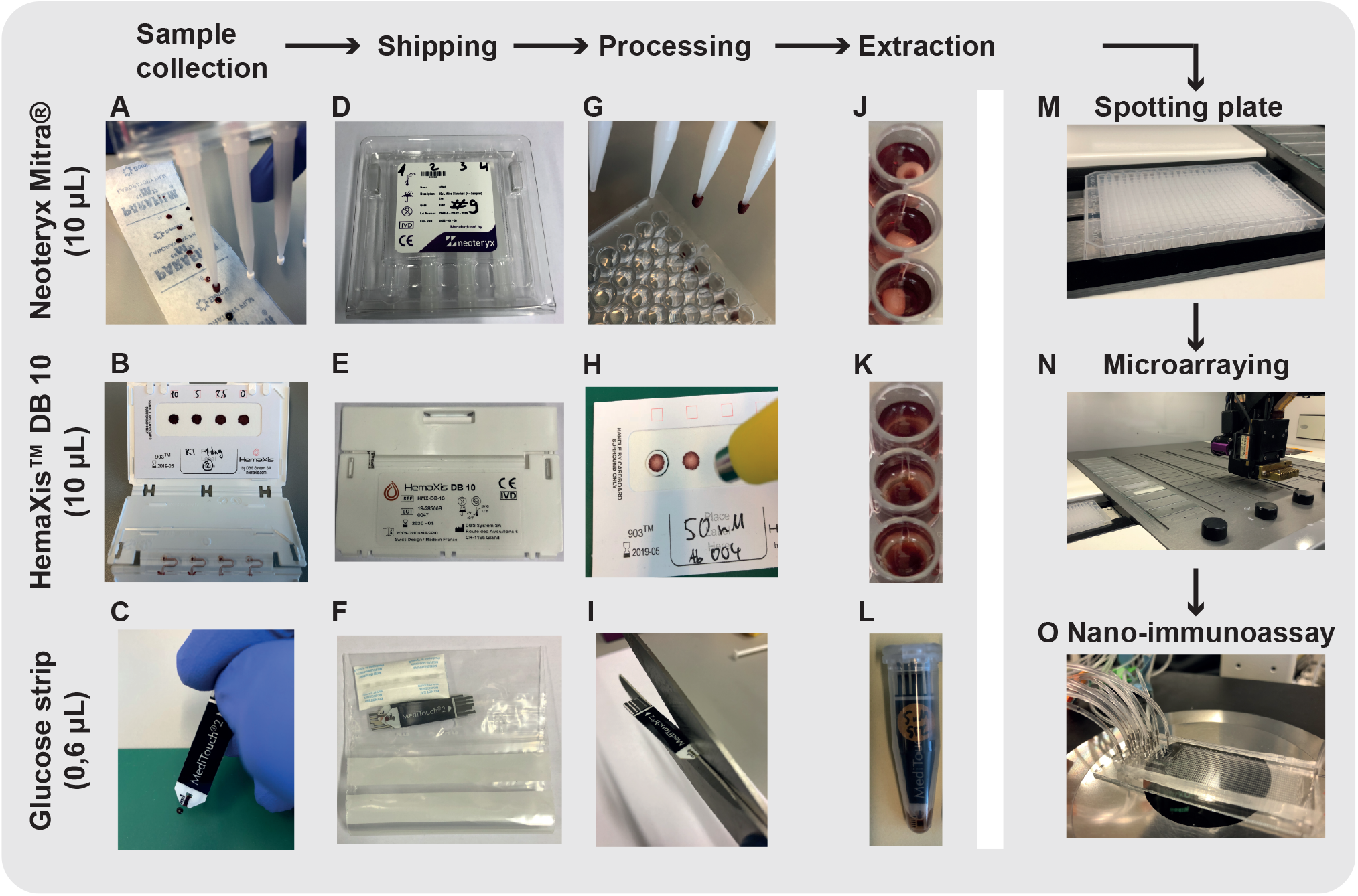
Ultra-low volume whole blood sampling and processing. Three devices were tested for ultralow volume whole blood sampling and extraction: Neoteryx Mitra®, DBS System SA HemaXis^™^ DB10, and glucose test strips. 10 *µ*L whole blood is collected by the **(A)** Mitra® d and **(B)** HemaXis^™^ DB10 devices. **(C)** 0.6 *µ*L whole blood is collected by the blood glucose test strip. **(D-F)** Blood samples are dried, allowing the devices to be shipped under ambient conditions by regular mail. **(G-I)** The devices are processed upon arrival at the laboratory. **(G)** Mitra® tips are removed and placed in a 96 well plate, **(H)** HemaXis^™^ DB10 cards are punched and the filter discs placed in a 96 well plate, **(I)** and the glucose test strip is cut to size and placed in an Eppendorf tube. **(J-L)** Blood samples are extracted in a buffer solution by overnight incubation at 4°C, followed by transfer to a spotting plate **(M)**. Samples are then microarrayed **(N)**, and analyzed with the NIA device **(O)**.

We evaluated the three collection methods by spiking human whole blood with different concentrations of anti-spike IgG and collected the blood with each collection device. We then extracted the dried blood from each device and spotted the samples for NIA analysis. We first tested an aqueous ethylenediaminetetraacetic acid (EDTA) solution and sonication for extraction [22], but the use of this buffer resulted in large and inconsistent spots during microarraying. We then tested PBS (phosphate-buffered saline, 1% BSA) or PBS (1% BSA, 0.5% Tween-20) [23] extraction buffers with overnight incubation at 4°C and found that the addition of 0.5% Tween-20 greatly improved the assay (data not shown). With this optimized sample extraction workflow we were able to quantitate anti-spike IgG from the dried blood samples (Fig. 4A-C, S7).

**Figure 4:**
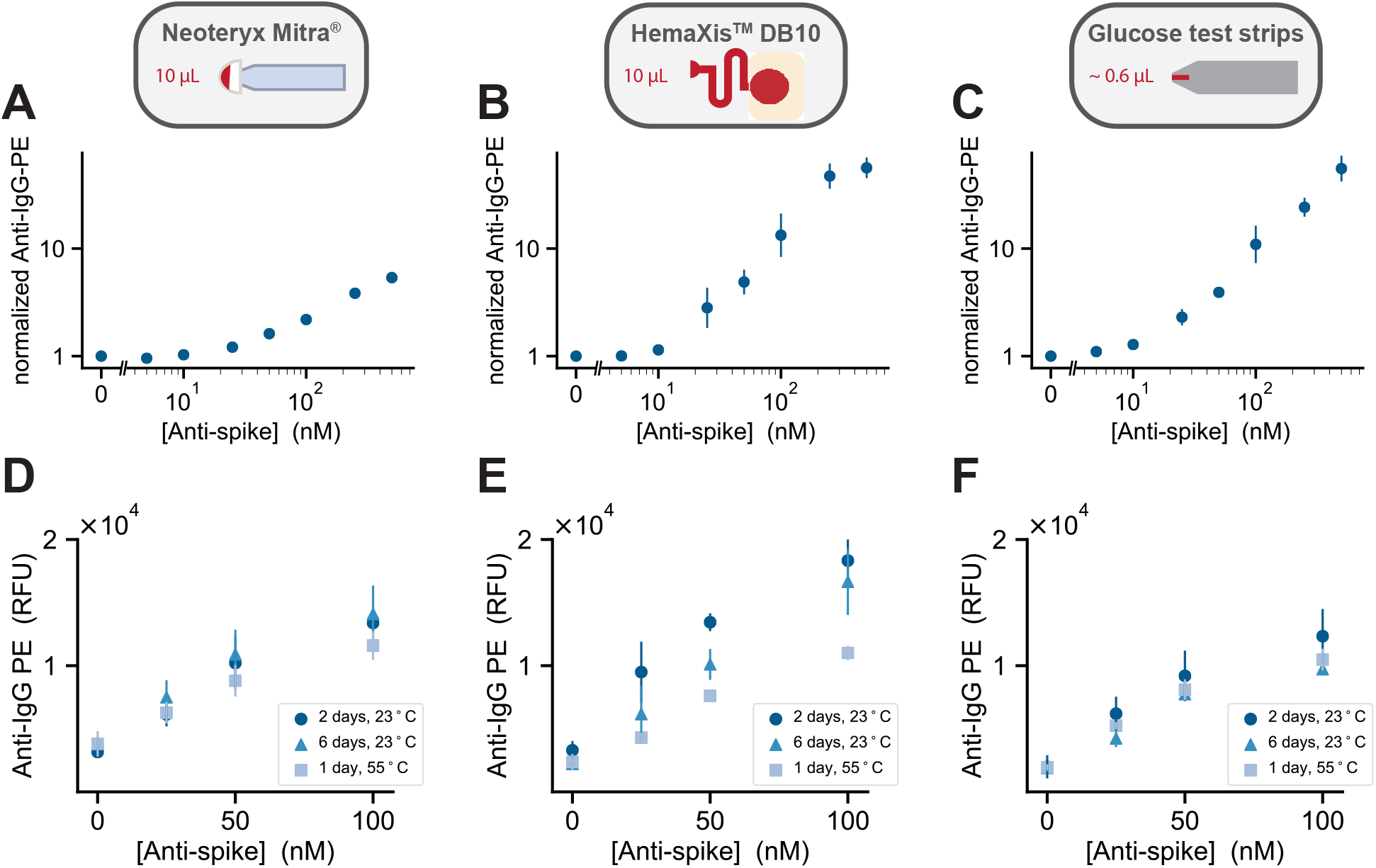
Ultra-low volume dried blood method characterization. The figure is organized into three columns each corresponding to the respective sampling method: Neoteryx Mitra^®^, HemaXis^™^ DB10, and glucose test strips. **(A-C)** Normalized NIA anti-IgG-PE signal versus concentration of anti-spike IgG present in dried whole blood samples collected with each of the three sampling methods. Data points represent means ±SD (n=4). The normalized signal is calculated by dividing the mean value for a given anti-spike concentration by the negative control mean value for 0 nM anti-spike. **(D-F)** Sample stability testing. NIA anti-IgG-PE measurements versus anti-spike IgG concentration. Blood samples were dried and then stored on each collection device for 1 day or 6 days at 23°C, and 1 day at 55°C.

To assess the variability of each sampling method we collected whole blood spiked with anti-spike IgG three separate times and compared the on-chip antibody signal (Fig. S8A-C). We calculated the coefficient of variation for the technical repeats for each anti-spike concentration tested and found that it did not exceed 15% for any of the three collection methods with an average CV of 5.7%, 7.7%, 9.2% for Mitra®, HemaXis™ DB10, and the glucose test strips (Fig. S8D). Although, there may be further variability introduced depending on how each individual collects his or her own blood sample, each device uses a hard-coded method to collect a specific volume of blood that leads to low variability for NIA antibody detection.

As we expect these blood sampling devices to be used decentralized, followed by shipping with regular mail to a central laboratory for analysis, an important factor to assess was sample stability. To determine stability of anti-spike IgG antibodies we allowed the blood to dry on each device followed by storage for two and six days at room temperature (23°C) before extraction and testing (Fig. 4D-F). Furthermore, depending on the climatic conditions, samples sent by post may be subject to higher temperatures, therefore we also stored the devices for 1 day at 55°C. For the Mitra® device we observed very little sample degradation after 6 days of storage at room temperature and slight sample degradation when stored at 55°C for one day. Very similar results were obtained for the glucose test strips. The HemaXis™DB10 devices were the most sensitive to prolonged and high temperature storage, but still resulted in sufficient signal for accurate quantitation.

Having established that ultra-low volume dried blood samples can be analyzed on the NIA platform, we tested the method with ultra-low volume whole blood patient samples collected with each of the three collection methods. As a surrogate for capillary blood we used 36 EDTA-whole blood samples from 21 RT-PCR confirmed COVID-19 patients and 15 presumed negative patients hospitalized for other reasons that served as negative controls. We collected whole blood samples in Geneva followed by shipping via regular mail to Lausanne for analysis. All positive samples are early seroconverts obtained within 14 days post diagnosis, and thus even standard, large-volume serum samples are challenging to analyze (Fig. 2C). For reference measurements we prepared plasma from EDTA-whole blood samples and performed S1 ELISA assays on the same patient samples.

We directly compared results obtained by ELISA performed on standard large-volume serum samples to NIA measurements performed on ultra-low volume dried whole blood samples collected with the three collection methods and processed as described above (Fig. 5). ELISA detected SARS-CoV-2 specific antibodies in 62% of all COVID-19 patient samples and found no detectable antibodies in any of the 15 presumed negative samples. For the three sampling methods we set the threshold between positive and negative calls to the intensity of the second highest negative sample. All three methods identified the same 62% anti-SARS-CoV-2 IgG positive samples as the reference ELISA, but the Mitra® method was able to detect antibodies in 33% additional RT-PCR positive samples and the HemaXis™ DB10 and glucose strip methods were able to detect an additional 29%. This proof of concept study demonstrates that ultra-volume whole blood samples can be collected and analyzed on the NIA platform. Surprisingly, even difficult to quantitate samples obtained within the first 14 days post onset of symptoms could be analyzed with this approach.

**Figure 5:**
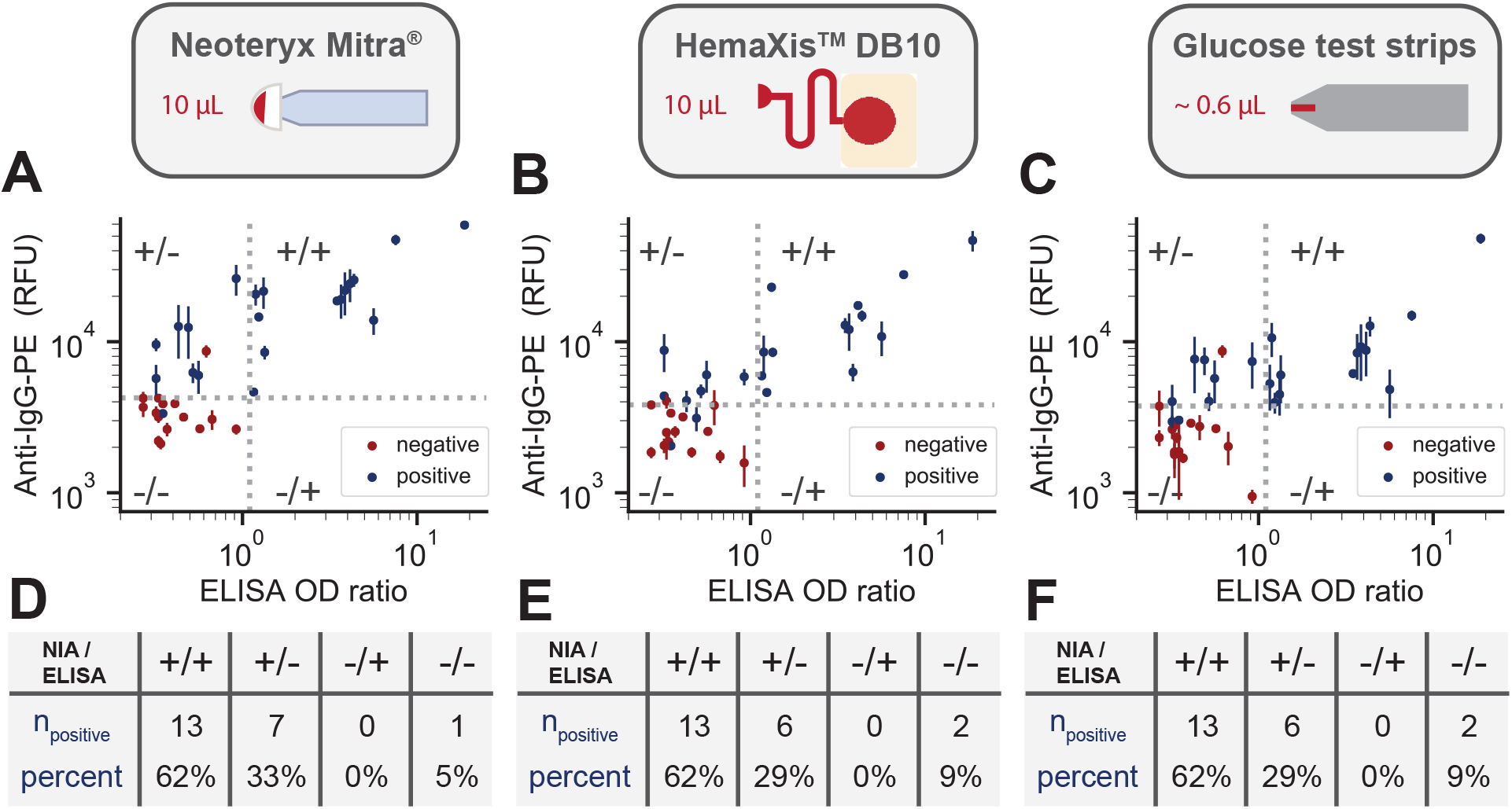
Ultra-low volume patient sample collection and analysis. **(A-C)** NIA anti-IgG-PE signal versus ELISA OD ratio for whole blood patient samples collected using each of the three sampling methods: Neoteryx Mitra®, DBS Systems SA HemaXis^™^ DB10, and glucose test strips. Data points are colored either blue or red corresponding to whether the samples were presumed positive or negative. The vertical dashed line represents the positive-negative cutoff for ELISA and the horizontal dashed line represents the chosen cutoff for the NIA measurements set equal to the 2nd highest negative measurement. Data points represent means ±SD (n=4). **(D-F)** Number of blue data points for each quadrant in plots **A-C**, respectively, along with the percentage of positive data points per quadrant.

## Discussion

We developed and validated a high-throughput nano-immunoassay device capable of analyzing 512-1024 samples in parallel. Detecting the presence of SARS-CoV-2 anti-spike IgG antibodies with a 512 sample throughput per device the method achieved a specificity of 100% and a sensitivity of 98% based on the analysis of serum samples from 155 positive SARS-CoV-2 infected and 134 negative individuals, performing as well or better than standard ELISA (Fig. 6A). When analyzing 1024 samples per device the method achieved a sensitivity of 95%. In addition to generating accurate binary classification of samples, NIA is also capable of returning accurate antibody titers.

**Figure 6:**
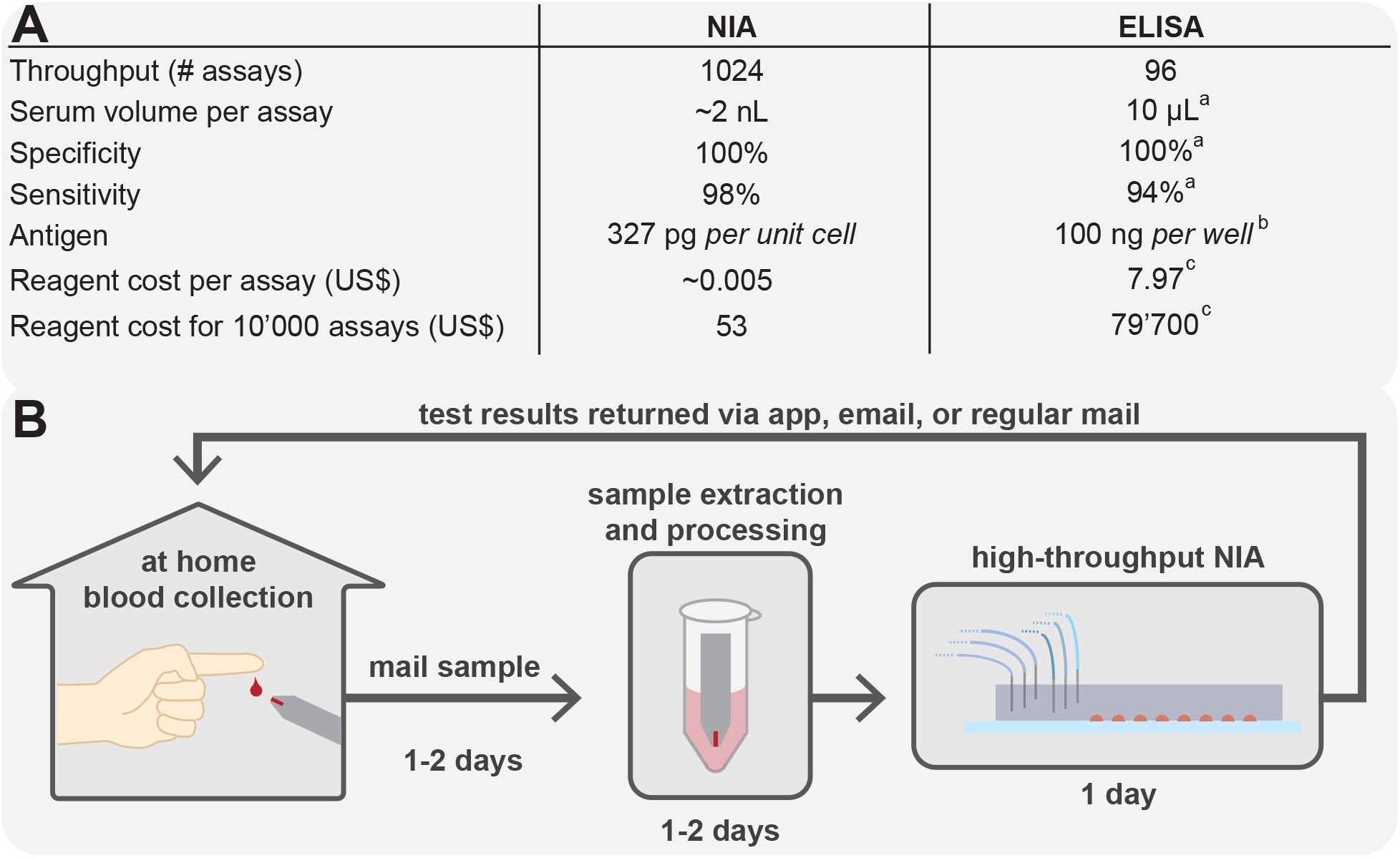
NIA performance table and conceptual home-based sample collection and centralized NIA analysis. **(A)** Comparison between the high-throughput NIA platform and standard ELISA. Sensitivity and specificity values are based on serum sample analysis for both NIA and ELISA. ^a^ based on EuroImmun ELISA kit (EI 2606-9601)[27], ^b^[23], ^c^ price based on abcam ELISA kit (ab274342). **(B)** A NIA based diagnostic workflow that uses decentralized ultra-low volume whole blood sample collection, shipping via regular mail, and centralized sample processing and analysis on a high-throughput NIA platform. Test results are analyzed and interpreted and then reported back via mobile app, email, or regular mail.

The method developed here is applicable to the large-scale characterization of serum samples collected as part of epidemiological studies, identify donors for plasma therapy, and vaccine trial support. A single researcher can achieve a throughput of 1-2 devices, or 512 - 1024 samples per day (analyzed in duplicate) in a small research laboratory not dedicated or equipped for high-throughput molecular diagnostics. Due to increases in efficiency, a small team of 3 can likely achieve a throughput of 6 devices or 3’072 samples per day (analyzed in duplicate). By comparison, the number of RT-PCR tests performed in all of Switzerland ranged from 6’000 to 18’000 tests per day between April and September 2020. Consumables, reagent consumption, and associated costs are negligible with NIA, which is an important consideration when compared to the high reagent cost of ELISAs and when considering potential reagent shortages that may be encountered during critical phases in a global pandemic.

To implement the method, laboratories require a commercially available contact microarrayer, and the ability to fabricate masks, molds and PDMS microfluidic devices. Instruments required for photolithographic fabrication of masks and molds are available at most major research universities, and mask fabrication can be outsourced to service-based companies. PDMS microfluidic device fabrication can be conducted in a standard research laboratory with low-cost equipment: a spin-coater, 80°C oven, and stereo-microscope. To run the microfluidic devices, standard pressure regulators, pressure gauges, and manual or solenoid valves are used. Device readout is performed on a standard epi-fluorescence microscope equipped with an automated stage.

Platform capabilities could be further expanded in the near term. We previously demonstrated that the platform can be used to perform multiplexed analysis, allowing 4 or more biomarkers to be tested for each sample [17, 24, 18]. This would allow the analysis of multiple antigens, cytokines, or inflammatory markers to provide insights into prior virus exposures and the response to infection. It should also be possible to detect IgA and IgM isotypes by using detection antibodies specific for these isotypes and to do so in a multiplexed format in which all 3 isotypes, IgG, IgA, and IgM are measured per sample. We also previously demonstrated that kinetic rate measurements can be performed in high-throughput with this method [25] and that digital ELISAs can be performed using the same technology in instances were lower limits of detection may be required [26, 11].

To enable large-scale studies we placed specific emphasis on the development of simple, low-cost, and ultra-low volume sample collection strategies and integration of these workflows with the nano-immunoassay platform. We characterized three blood sampling devices and tested all of them with patient samples. Two of these methods are commercially available devices: Mitra® from Neoteryx and HemaXis™ DB10 from DBS System SA. We also explored the possibility of repurposing low-cost, and readily available blood glucose test strips for blood sample collection and shipment. The two commercial devices allow the collection of a minimal volume of 10 *µ*L whereas the blood glucose test strip collects as little as 0.6 *µ*L whole blood. Such small volumes allow untrained personnel to fingerprick and collect whole blood, eliminating the need for phlebotomists and the inconvenience of visiting a hospital or point of care location. The collected blood is allowed to dry on the devices, and we could show that these dried blood samples can be analyzed after 6 days of storage at ambient temperatures eliminating the need for a cold chain.

The combination of a high-throughput, highly specific and sensitive nano-immunoassay, and the ability to analyze minute volumes of dried blood samples has enormous potential for SARS-CoV-2 serology, epidemiological studies, vaccine trial and therapeutic development support. Further areas of use could be large-scale seroprevalence studies in low- and middle-income countries [28] without sufficient in-country laboratory capacity by sending specimens by international mail. Especially in the current SARS-CoV-2 pandemic, population-based seroprevalence studies could elucidate some urgent questions such as the impact of COVID-19 in Africa [29].

New technology and method developments such as those reported here will make it possible to overcome the current centralized molecular diagnostics paradigm, which is focused on hospitals and point of care settings rather than on patients and individuals seeking simple, affordable, and convenient molecular diagnostics. The need to visit a clinic is an inconvenience for everyone and can be an insurmountable obstacle for many. The requirement for venipuncture blood collection, sample pre-treatment, and expensive ELISAs prohibits broad testing and contributes to high health care costs. NIA makes it possible for individuals to purchase a simple blood sampling kit containing a lancet, a blood sampling device, and a return mail envelope, at a local pharmacy or supermarket (Fig. 6B). The kit can be used easily and conveniently in the privacy of one’s own home, where a simple fingerprick is made and the blood collected by the device. The device with the collected blood can then be sent without special biosafety requirements by regular mail to a central laboratory which analyzes the blood sample for one or more biomarkers, interprets the data, and returns the test results to the individual via smart phone, email, or regular mail. Furthermore, each blood sample is sufficient to conduct many molecular diagnostic assays with NIA. The whole process from kit purchase to results could take less than a week, which is fast enough for a vast majority of tests that are not particularly time critical. Decentralized and simple sample collection coupled with centralized, next-generation high-performance molecular tests will broaden access to molecular diagnostics, and increase the use of testing. During a global pandemic, such technologies could enable the collection of critical epidemiological data, provide instrumental data for vaccine development, and provide information to individuals on their health status.

## Methods

### Microfluidic chip fabrication

The designs for the flow and control layer of the device were drawn with AutoCAD software and are available for download (lbnc.epfl.ch), we then used standard photolithography to fabricate the molds for each layer. Three replicates of the design are fitted onto a 4 inch silicon wafer so that three devices can be made in a single fabrication process. SU-8 negative photoresist was used to create the control channel features (GM 1070, Gersteltec Sarl) with a height of 30 *µ*m, while AZ 10XT-60 positive photoresist (Microchemicals GmbH) was used to generate flow channel features with a height of 15 *µ*m. After development, the flow layer mold was annealed at 180°C in a convection oven for two hours to obtain rounded features. Afterwards each of the wafers was treated with TMCS (trimethylchlorosilane) and coated with PDMS (Sylgard 184, Dow Corning). For the control layer ∼50 g of PDMS with an elastomer to crosslinker ratio of 5:1 was prepared, whereas for the flow layer a 20:1 ratio of elastomer to crosslinker was spin coated at 400 rcf to yield a height of ∼50 *µ*m. Both PDMS coated wafers were then partially cured for 20 minutes at 80°C, after which devices from the control layer were cut out and the inlets for each control line were punched (OD = 889 mm) using a precision manual-punching machine (Syneo, USA). Each control layer is then aligned onto the flow layer by hand using a Nikon stereo microscope. The aligned devices were then placed at 80°C for 90 minutes, allowing the two layers to bond together so that the entire device can then be cut and removed from the flow wafer. After that, the flow layer inlets were punched.

### Immunoassay reagents

For our on-chip immunoassay we used biotinylated mouse anti-His antibodies (Qiagen, 34440) to immobilize His-tagged SARS-CoV-2 antigens on the surface of our assay chambers. The prefusion ectodomain of the SARS-CoV-2 spike glycoprotein (the construct was a generous gift from Prof. Jason McLellan, University of Texas, Austin [30]) was transiently transfected into suspension-adapted HEK293 cells (Thermo Fisher) with PEI MAX (Transfection grade linear polyethylenimine hydrochloride, Polysciences) in Excell293 medium. Incubation with agitation was performed at 37°C and 4.5% CO2 for 5 days. The clarified supernatant was loaded onto Fastback Ni2+ Advance resin column (Protein Ark) eluted with 500 mM imidazole, pH 7.5 in PBS. For proof-of-concept experiments, chimeric anti-spike antibodies were purchased from Sino Biological (40150-D002, 40150-D003, 40150-D004, 40150-D005). We spiked the chimeric anti-spike antibodies into human serum (Sigma-Aldrich, H4522) and whole blood (ZenBio, SER-WB). For detecting human IgG, we used PE labeled goat anti-Human-IgG (Abcam, ab131612).

### Sample preparation

#### Serum

Serum samples were collected from 155 RT-PCR confirmed COVID-19 patients and 134 negative control sera obtained in 2013/14 and 2018 before the start of the pandemic (including 50 children). All samples were collected according to the local ethical guidelines and ethical approval was waived by the ethics committee of the University Hospital of Geneva (HUG). We used days post onset of symptoms (dpos) according to patient history or days post diagnosis (dpd) in case dpos was unknown. All samples were stored at -20°C until analysis. In order to handle patient serum samples in a Biosafety Level 1 laboratory, patient serum samples were heat treated at 56°C for 30 minutes and Triton X-100 (Fisher Scientific) was added to the samples to a final concentration of 1%. In order to optimize spotting parameters and the on-chip immunoassay, proof-of-concept experiments involving human serum spiked with chimeric anti-spike antibodies were also carried out with the addition of Triton X-100. For dilution series experiments, patient serum samples were diluted in a PBS solution containing 2% BSA. Additionally, 10 *µ*M fluorescein isothiocyanate(FITC)-dextran (10 kDa) was added as a tracer to each sample in order to assess whether similar volumes of serum samples were spotted (Fig. S9).

### Whole and dried blood

Human Whole Blood - Frozen, 10ml was obtained from AMS Biotechnology (Europe). Anti-spike antibodies (Sino Biological) were added to whole blood to the desired concentration from a stock solution of 2 *µ*M. To simulate a fingerprick collection, 15-20 *µ*L of whole blood samples were pipetted on parafilm and then collected on the sampling device where 10 *µ*L were collected with HemaXis^™^ DB10 (DBS System SA) or Mitra® Clamshell (Neoteryx). The samples were dried and then stored in their original container without further protection. Samples were kept at room temperature for 1, 2 or 6 days, or in a 55°C oven for 1 day. The dried blood stored on HemaXis^™^ filter paper were cut using an 8 mm biopsy puncher and scalpel and the tip of the Mitra® devices were removed with tweezers. The samples were placed in a 96-well plate and filled with 200 *µ*L of cold extraction buffer (1xPBS, 1% BSA, 0,5% Tween20) [23] and incubated overnight at 4°C with 300 rpm agitation. Around 150 *µ*L per well of the supernatant was recovered and stored at -20°C until analysis. Alternatively, a small volume of blood (around 0.6 *µ*L) was collected with a Meditouch 2 glucose test strip (Medisana), dried and stored in a box kept at room temperature for 1,2 or 6 days, or in a 55°C oven for 1 day. Extraction was performed by placing the strip at the bottom of a 1,5 mL tube filled with 30 *µ*L of extraction buffer overnight at 4°C.

As a substitute for capillary blood taken from a fingerprick, we used leftover EDTA whole blood samples drawn from hospitalised COVID-19 patients at different time points post diagnosis for routine clinical laboratory analysis. EDTA whole blood samples were stored at 4°C for a maximum of seven days before they were applied to the dried blood collection devices. 20 *µ*L of EDTA whole blood was pipetted on a parafilm sheet, and collected with a Mitra® device or glucose strip. For the HemaXis^™^ device, 10 *µ*L of whole blood was directly pipetted on the filter card. The samples were dried for 30 minutes and placed in a plastic bag with silica gel before shipping. The samples were extracted as described above 5 days after collection and shipping, microarray spotted on a glass slide on day 6 and the nano-immunoassay chip was run on day 7. To determine antibodies against SARS-CoV-2 S1 protein by Euroimmun S1 ELISA, EDTA whole blood samples where centrifuged for 5min at 1200x g at 4°C and 200 *µ*L of plasma were stored at 4°C until analysis.

### SARS-CoV-2 serological assays

Euroimmun S1 IgG ELISA (Euroimmun AG, Lübeck, Germany, # EI 2606-9601 G) was performed according to the manufacturers instructions and was run on a Dynex Agility (RUWAG Handels AG, Bettlach, Switzerland). OD ratios were calculated by dividing the OD450 of each sample by the OD450 of a calibrator that was run on each plate. Results equal or above OD ratio of 1.1 were considered positive. The LIAISON SARS-CoV-2 S1/S2 IgG ELISA was run on the LIAISONR XL analyzer, (Diasorin, Italy), the EDI Novel Coronavirus COVID-19 IgG ELISA (Epitope Diagnostics, USA) on the DSX analyzer (Dynex, Switzerland) and the Elecsys Anti-SARS-CoV-2 N (anti-N total antibodies) as well as the Elecsys Anti-SARS-CoV-2 S (anti-S1-RBD total antibodies) on the Cobas e801 analyzer (Roche Diagnostics, Switzerland) according to the manufacturers instructions.

### Microarray spotting

25 *µ*L of each sample were loaded into a 384 microwell plate (ArrayIt, MMP384). An MP3 microarray printing pin (Arrayit) was used to spot the samples onto an epoxy-coated glass slide using a QArray2 microarrayer (Genetix). The presence of Triton X-100 in the serum samples had a significant effect on the spot diameter. To account for this we increased the dimensions of the spotting chamber and set the inking and stamping time to 50 ms and 1 ms, respectively. In addition, it was critical that the ambient humidity was below ∼42%, otherwise the spots would become too large and merge together. After spotting, the PDMS chip was aligned on top of the sample spots using a stereo microscope and bonded over night at 40 °C.

### Running on-chip immunoassays

Control lines were filled with PBS, attached to the chip and pressurized at 145 kPa. The NIA device is a low-complexity device, and can therefore be either regulated with simple manual 3 way toggle switch valves or computer controlled solenoid valves (only needed in order to make the entire on-chip workflow fully automated). Control and flow pressures are set by standard pressure regulators using a building air supply as the pressure source and monitored with two pressure gauges, respectively. Detailed descriptions of a standard MITOMI setup [31] and more sophisticated computerized microfluidic control setups for controlling complex multi-layer microfluidic devices have been previously published [32, 33]. While isolating the spotted sample with the neck valve closed, the lower half of the unit cells were patterned with BSA-biotin (Thermo Fisher, 29130) and neutrAvidin (Thermo Fisher, 3100). First BSA-biotin was flowed at aconcentration of 2 mg/mL for 20 minutes, then neutrAvidin was flowed at a concentration of 1 mg/mL for 20 minutes. A flow pressure of 27-34 kPa was maintained for each of the flow steps. Afterwards the button valve was actuated and BSA-biotin was flowed for an additional 20 minutes. In between each of these steps a solution of 0.005% Tween 20 (Sigma, P1379) in PBS was flowed for 5 minutes to wash away any unbound material. After surface functionalization, a 1 *µ*g/mL solution of biotinylated anti-His antibody in 2% BSA in PBS was flowed for 20 minutes. Next a 6.7 *µ*g/mL solution of His-tagged SARS-CoV-2 spike protein in 2% BSA in PBS was flowed for 20 minutes. The spike protein solution contained 25% chicken serum (Sigma, C5405), which served to block the surface and reduce non-specific binding. Before each of these steps the solution was first flowed for 2 minutes with the button valves down, allowing the solution to flow evenly into the entire device. After flowing each solution for 20 minutes, a wash step was performed by flowing 0.005% Tween PBS for 5 minutes with the button valves down. Once the spike protein was attached to the surface, the serum spots were re-solubilized by opening the neck valve and allowing 0.005% Tween PBS to flow into the spotting chamber while the outlet valve was closed. The neck valve was then closed and 0.005% Tween PBS was flowed for 5 minutes to prevent cross-contamination of samples in neighboring unit cells. The sandwich valves were then closed and the neck valve was released to allow any antibodies present in the serum spot to diffuse into the assay chamber. After an incubation of 70 minutes the button valve was opened and a second incubation of 60 minutes was carried out, permitting any anti-spike antibodies to bind to the spike protein. Any unbound material could then be washed away by flowing 0.005% Tween PBS for 5 minutes with the button and neck valves closed. A 5.6 *µ*g/mL solution of anti-IgG-PE was then flowed for 2 minutes with the buttons down, then for 10 minutes with the buttons up. The buttons were then closed and any unbound detection antibody was washed away by flowing 0.005% Tween PBS for 5 minutes. Each unit cell was then imaged with an exposure time of 300 ms using a Nikon ECLIPSE Ti microscope equipped with a LED Fluorescent Excitation System, a Cy3 filter set, and a Hamamatsu ORCA-Flash4.0 camera (C11440).

## Data Availability

All data referred to in the manuscript is available on MedRxiv as supplemental files.

http://lbnc.epfl.ch/resources.html

## Data analysis

The detection antibody signal was quantified using a custom Python script. ROC analysis was performed using GraphPad Prism. Cutoff values for the NIA measurements were chosen by maximizing both the specificity and sensitivity values. *EC*_50_ values were determined by fitting the dilution series data (1:256 - 1:8) for each sample to a saturation binding curve,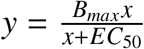. For all samples *B*_*max*_ was set to 65000.

## Acknowledgments

We would like to thank the Protein Production and Structure Core Facility team at EPFL, specifically Dr. David Hacker, Dr. Florence Pojer, Dr. Kelvin Lau, Laurence Durrer and Soraya Quinche for production and purification of Spike 2P in mammalian cells. We thank Neeraj Dhar and the Mckinney lab at EPFL for their help in sample inactivation in BSL-3. We thank Isabelle Arm-Vernez and Sabine Yerly for technical assistance as well as Barbara Lemaitre and Catia Machado Delgado for sample preparation. In addition, we thank Guiseppe Togni for running serological assays. We kindly thank DBS System SA for generously providing us with HemaXis^™^ DB10 devices. This work was supported by the European Research Council under the European Union’s Horizon 2020 research and innovation program Grant 723106 (SJM), SNF Project grant 182019 (SJM), SNF NRP 78 Covid-19 grant 198412 (SJM, IE, and BM), EPFL (SJM) and a grant from the Private Foundation of the Geneva University Hospital (IE).

## Author contributions

SJM, BM, and IE developed the project. PC, DOA, NV, LK provided resources and advice on experiments. ZS, GM, HMY, performed experiments. ZS, GM, HMY, IE, BM, SJM, designed experiments, analyzed data, and wrote the manuscript.

### Competing interests

The authors declare no conflict of interest.

## Supplementary Figures

**Figure S1:**
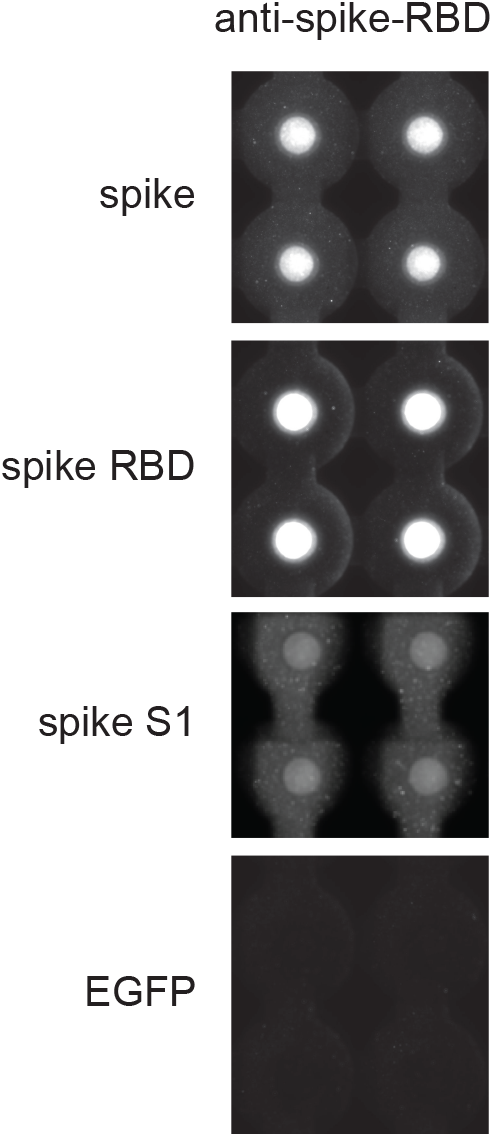
Evaluation of SARS-CoV-2 antigens. NIA images showing anti-IgG-PE signals obtained when spike, RBD and S1 antigens in combination with an anti-S1 primary antibody were tested. EGFP served as a negative control.

**Figure S2:**
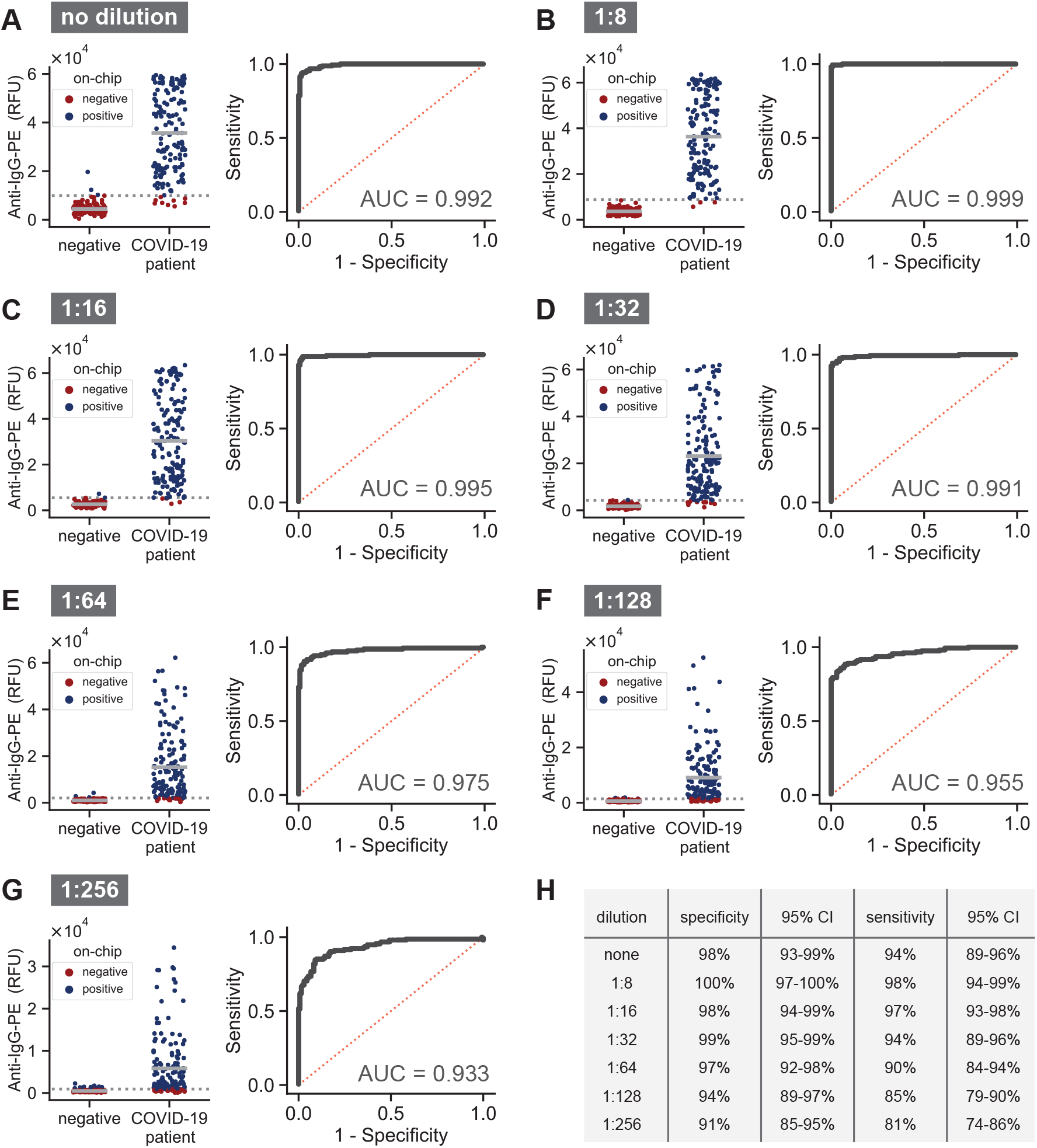
NIA measurements for a range of sample dilutions. **(A-G)** NIA measurments for different dilutions of patient serum samples categorized according to whether the sample was pre-pandemic negative or from COVID-19 patients. Data points represent mean values (*n* = 3). Corresponding ROC curves are shown to the right of the plotted data. **(H)** Specificity and sensitivity values calculated for each dilution according the dashed cutoff line shown in plots **A-G**.

**Figure S3:**
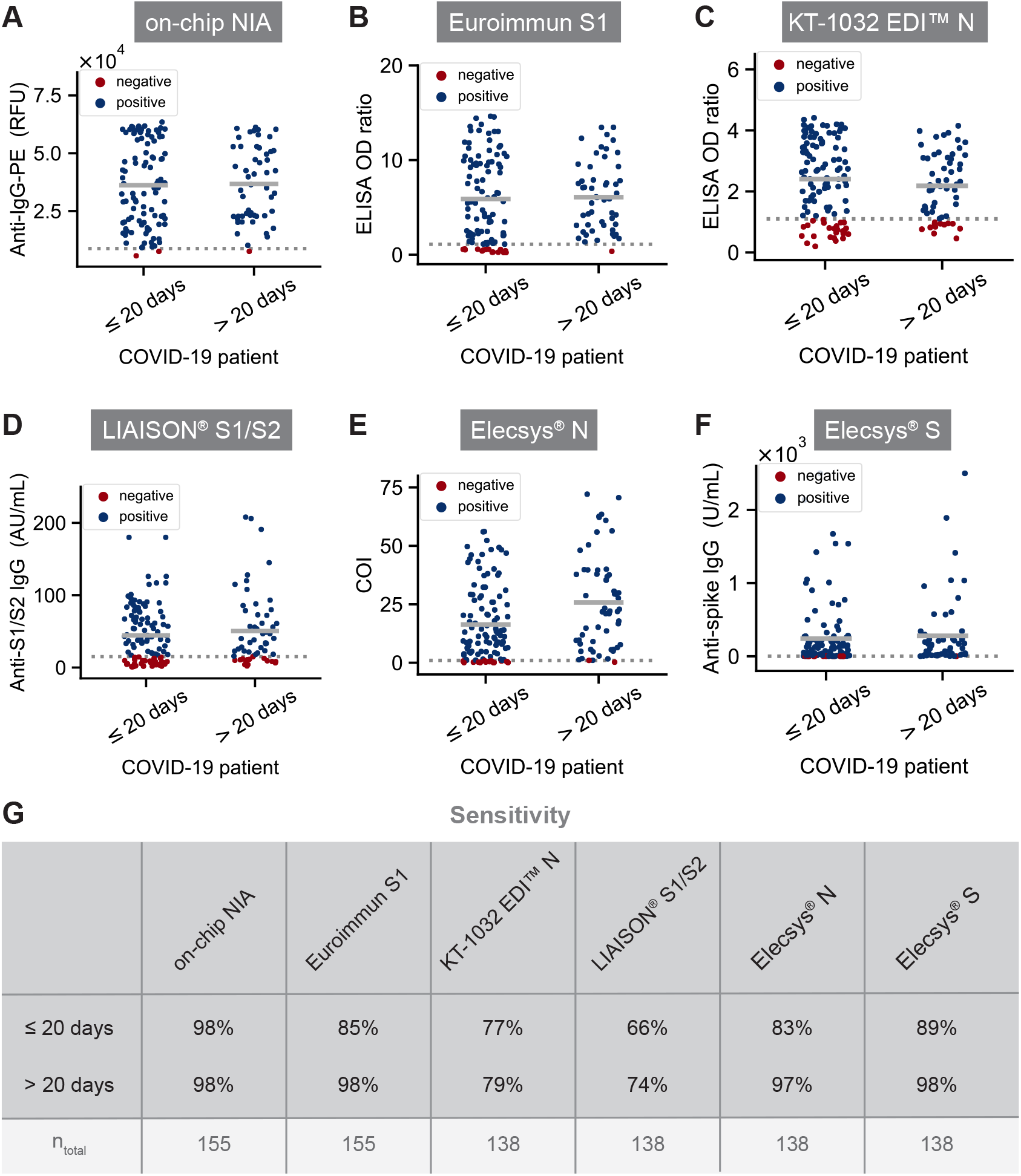
Comparison of NIA with commercial assays. **(A-F)** Levels of anti-SARS-CoV-2 IgG antibodies present in serum collected from COVID-19 patients ≤20 days or *>*20 days post onset. **(G)** Sensitivity values calculated for each assay according to the dashed cutoff line shown in plots **A-F**. The manufacturer recommended cutoffs were used for the commercial assays. The total number of patient serum samples tested for each assay is listed below the sensitivity values.

**Figure S4:**
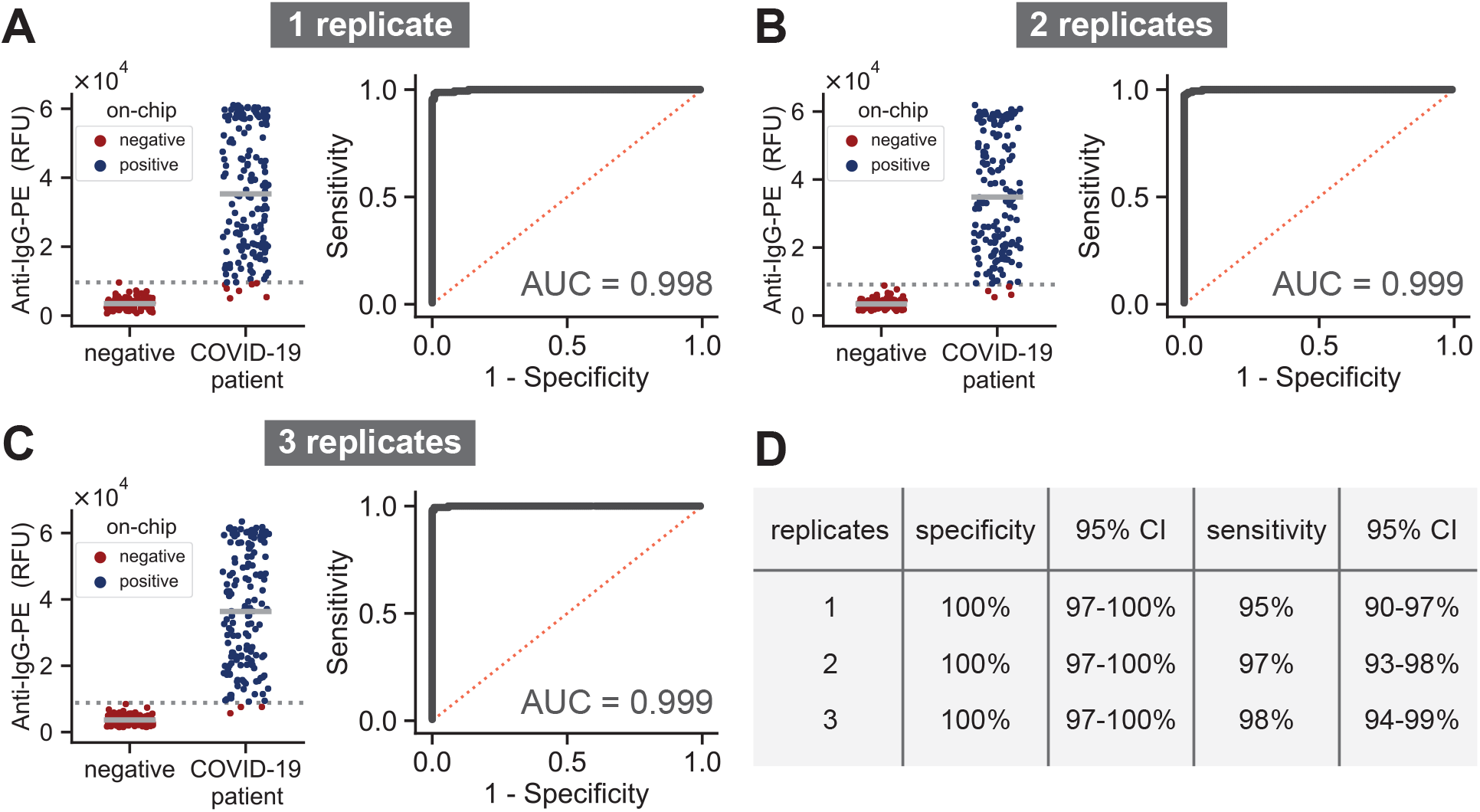
NIA replicates. **(A-C)** Mean anti-IgG-PE signal for one, two or three on-chip replicates shown for a 1:8 serum dilution, along with the corresponding ROC curves. **(D)** Specificity and sensitivity values calculated according number of on-chip replicates and based on the dashed cutoff line shown in plots **A-C**.

**Figure S5:**
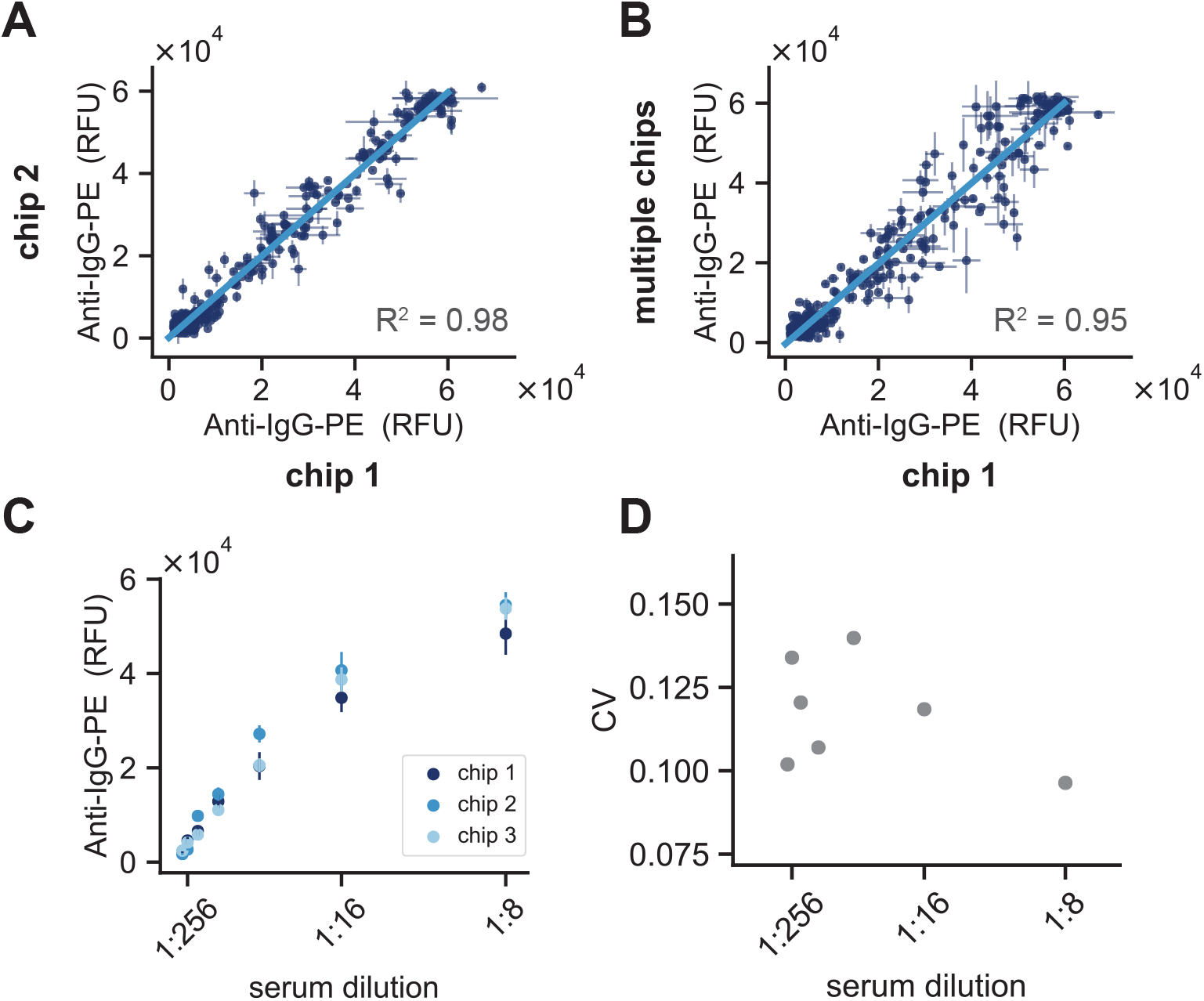
Device-to-device variation. **(A)** Correlation of anti-IgG-PE signals obtained from two separate chips that were prepared using the same 1:8 serum sample dilutions. **(B)** Anti-IgG-PE measurements collected from a total of 6 chips versus measurements for the same samples collected on a single chip. Sample dilutions were prepared separately for each of the chips. **(C)** NIA measurements for a reference serum dilution series measured on three separate chips. **(D)** Coefficient of variation calculated for the three measurements of each reference serum dilution.

**Figure S6:**
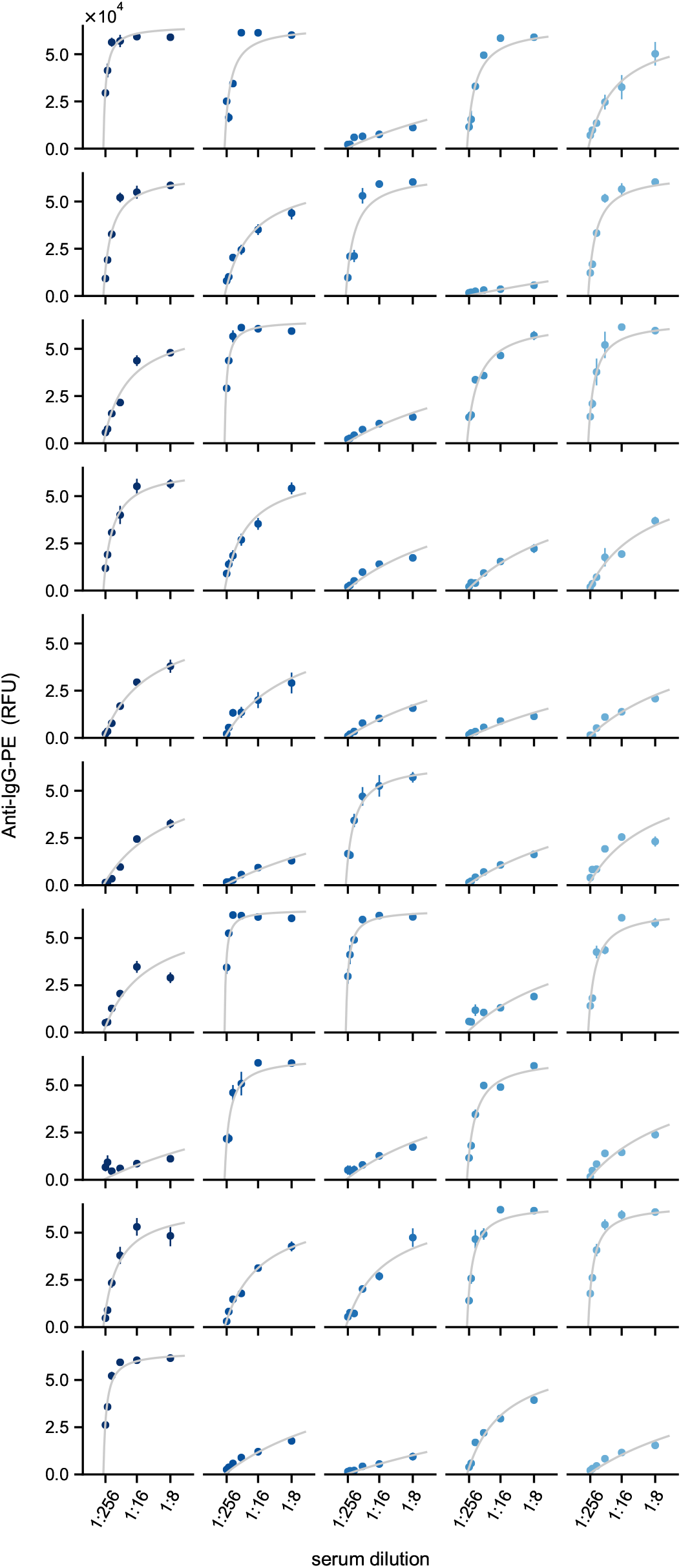
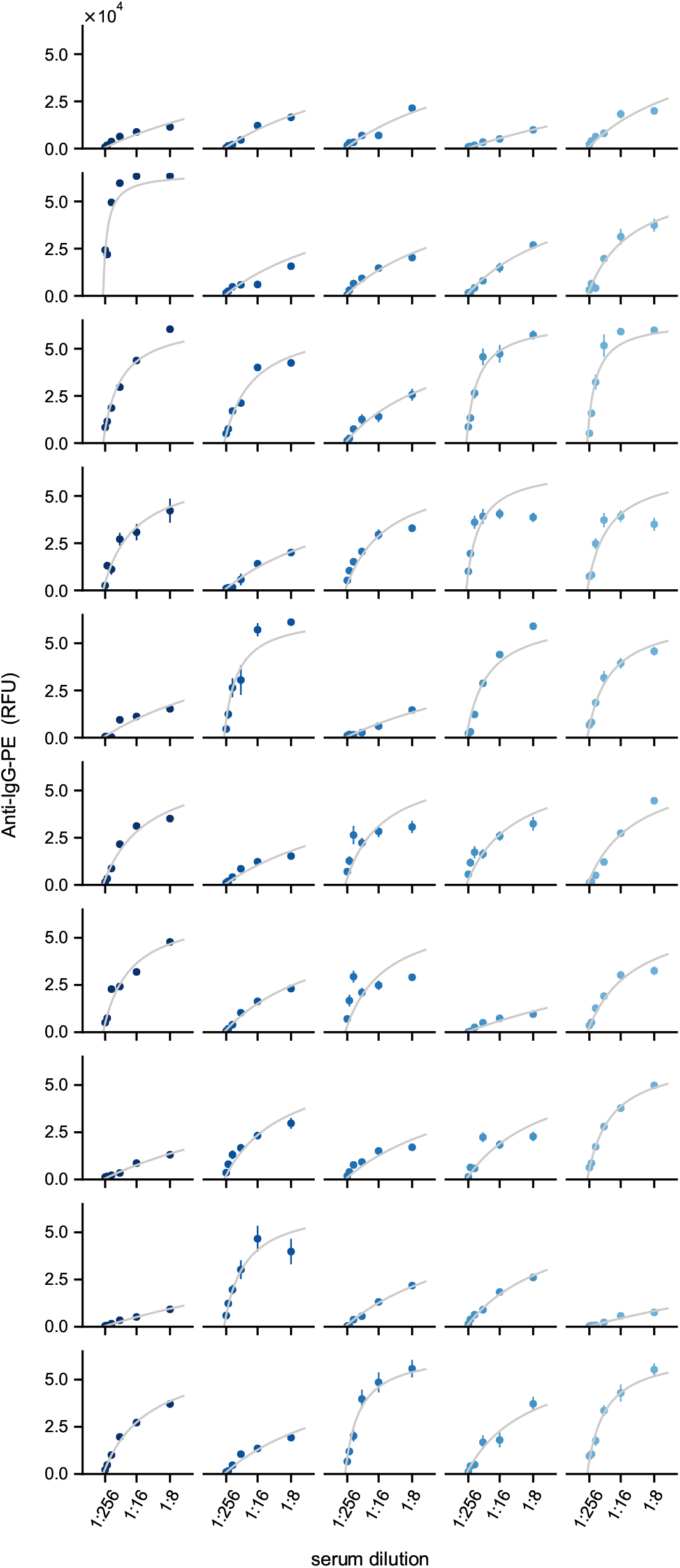
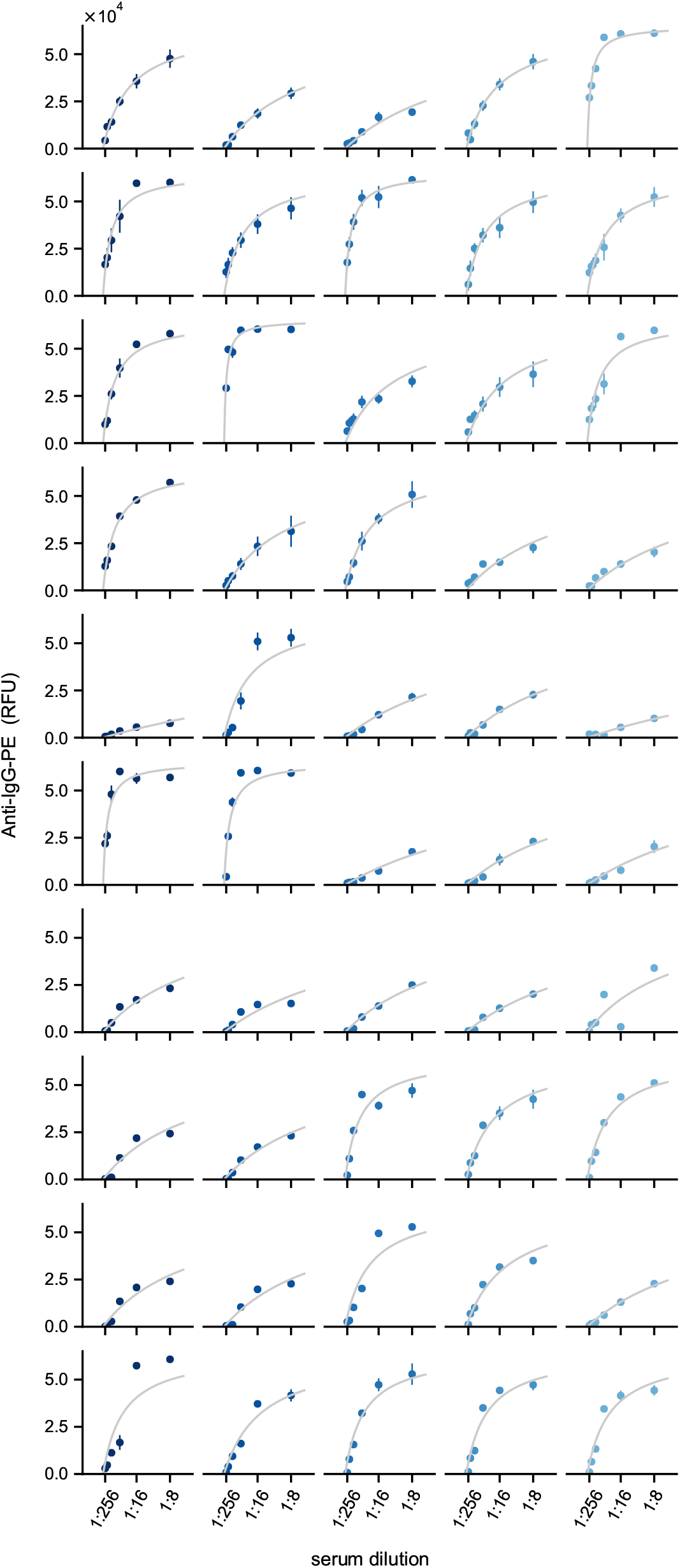
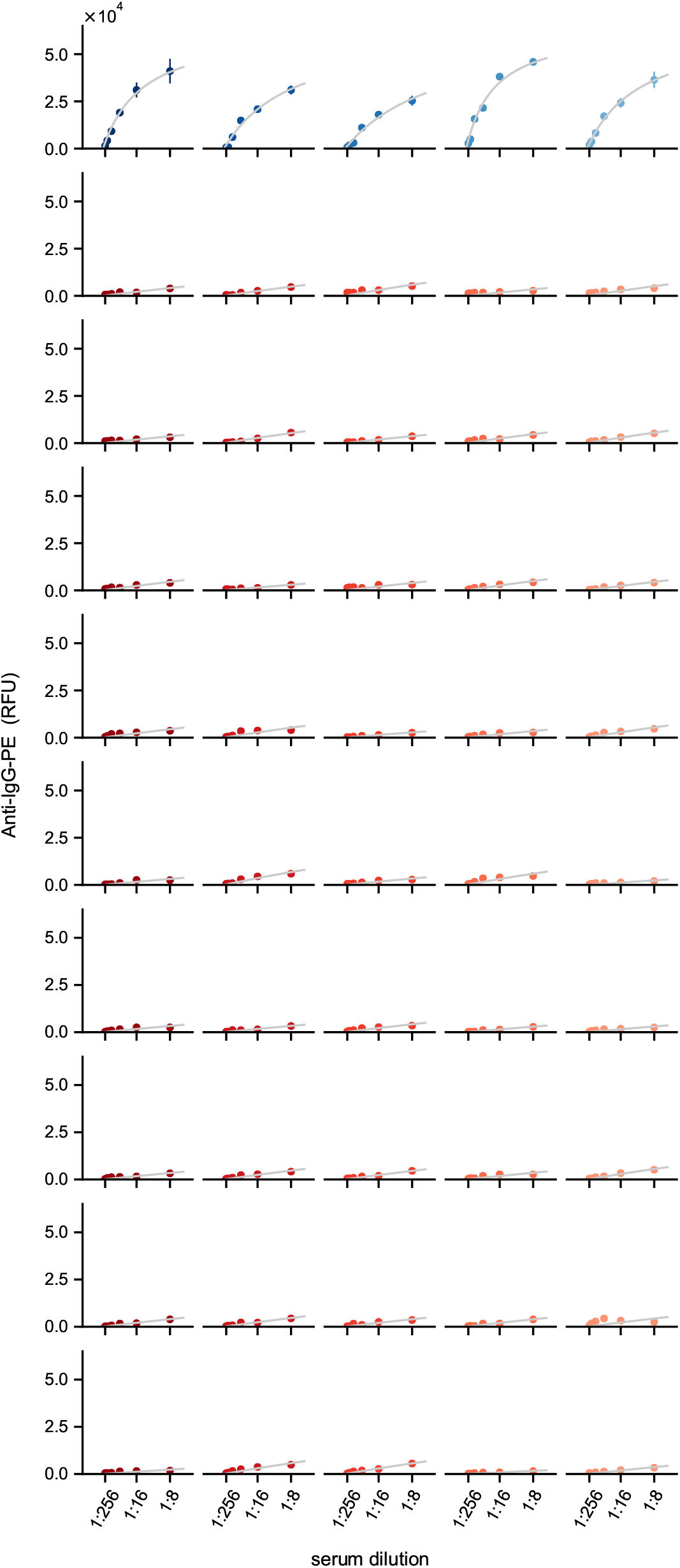
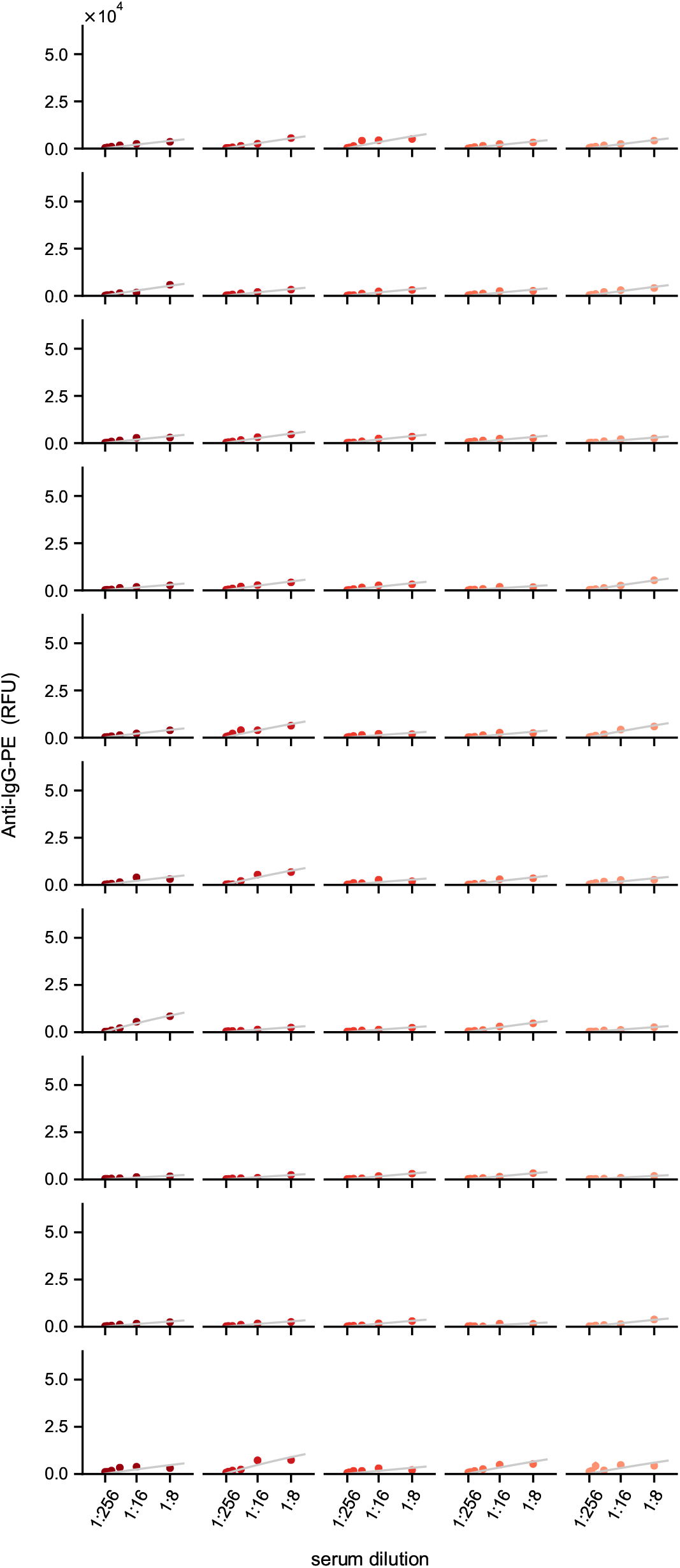
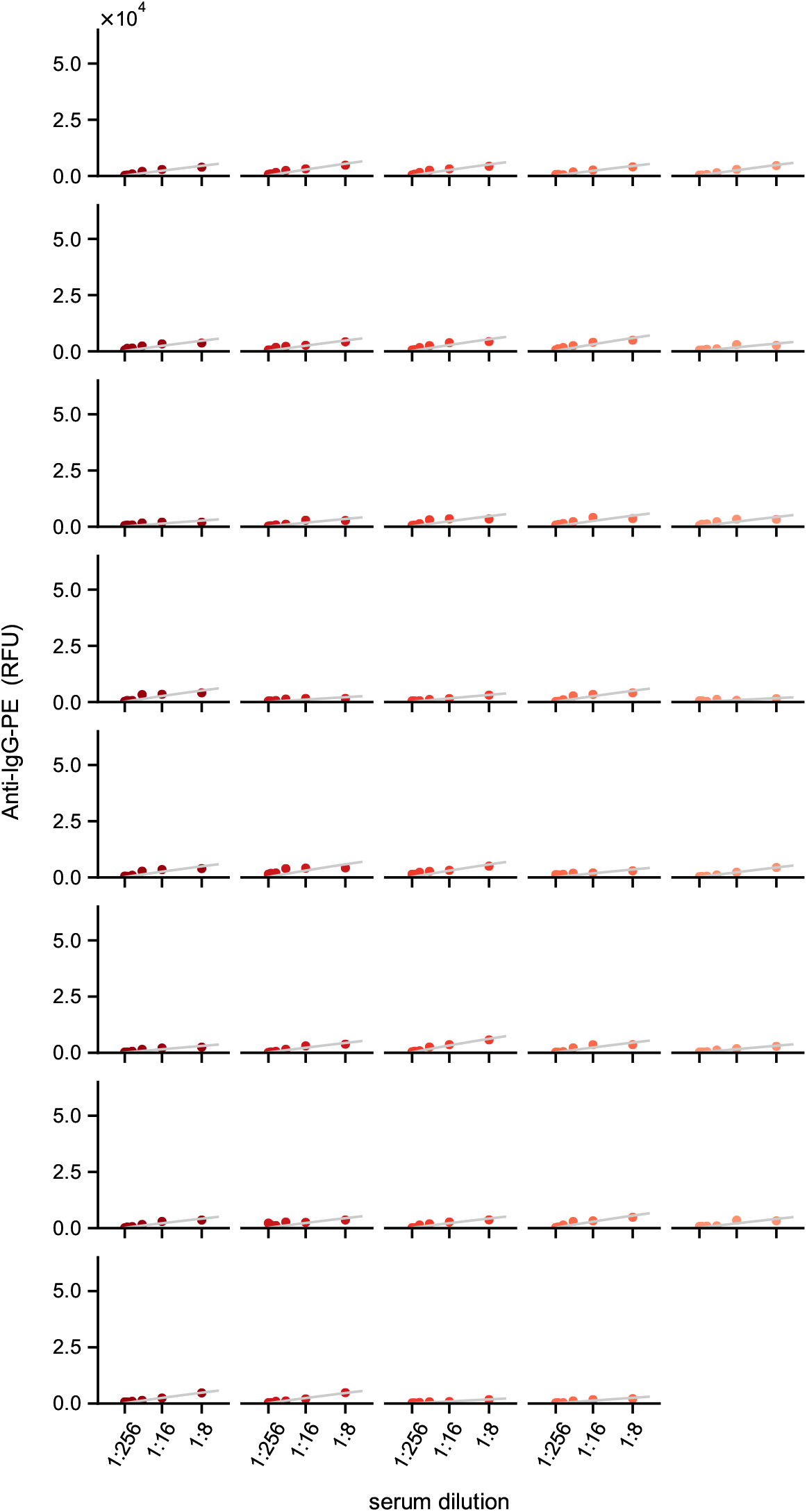
Complete serum dilution data. Data points are colored blue or red corresponding to dilutions from negative or positive patient serum samples, respectively.

**Figure S7:**
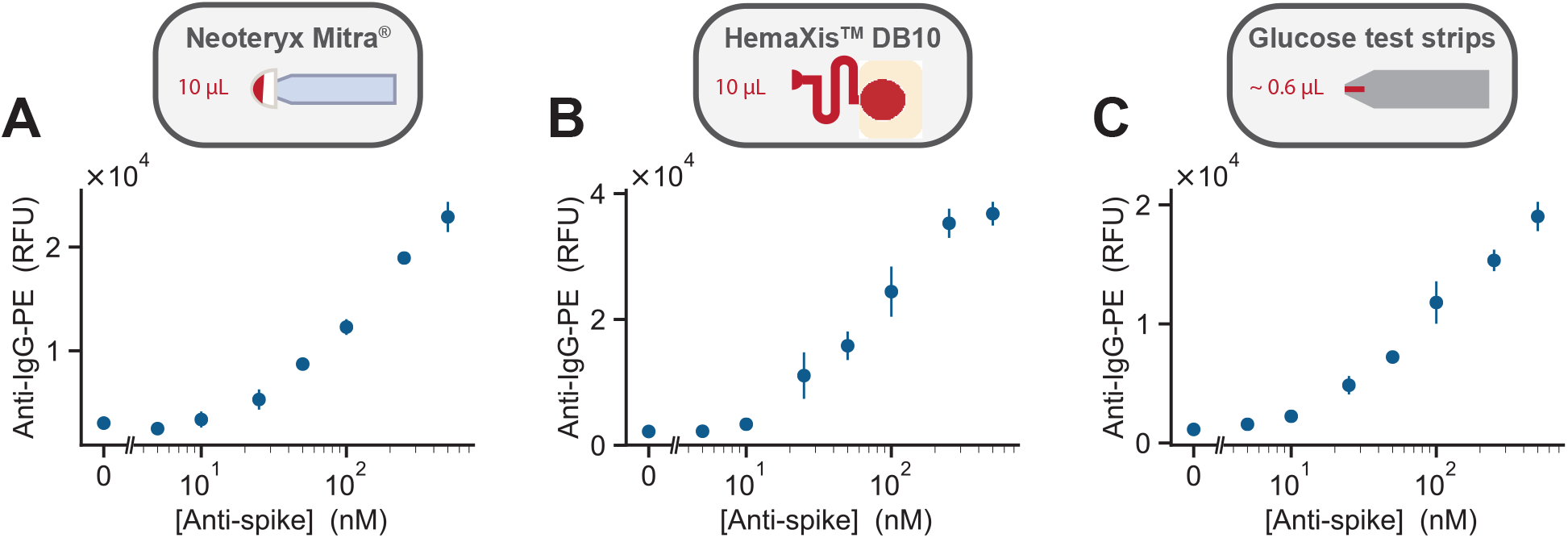
Detection of anti-spike IgG in whole blood. **(A-C)** Non-normalized on-chip anti-IgG-PE signal versus the concentration of anti-spike-IgG in whole blood sampled using each of the three methods: Mitra^®^, HemaXis^™^ DB10, and glucose test strips (shown in this order from left to right).

**Figure S8:**
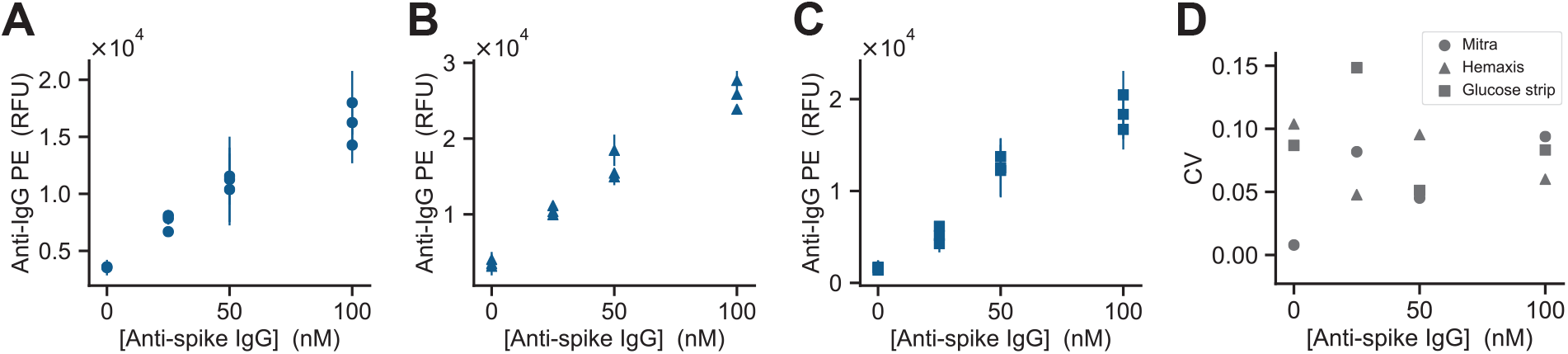
Technical replicates for ultra-low volume whole blood sampling methods. **(A-C)** On-chip anti-IgG-PE signal versus the concentration of anti-spike-IgG in whole blood for three technical replicates sampled using each of the three methods: Mitra®, HemaXis^™^ DB10, and glucose test strips (shown in this order from left to right). **(D)** Coefficient of variation versus the concentration of anti-spike-IgG for each blood sampling method.

**Figure S9:**
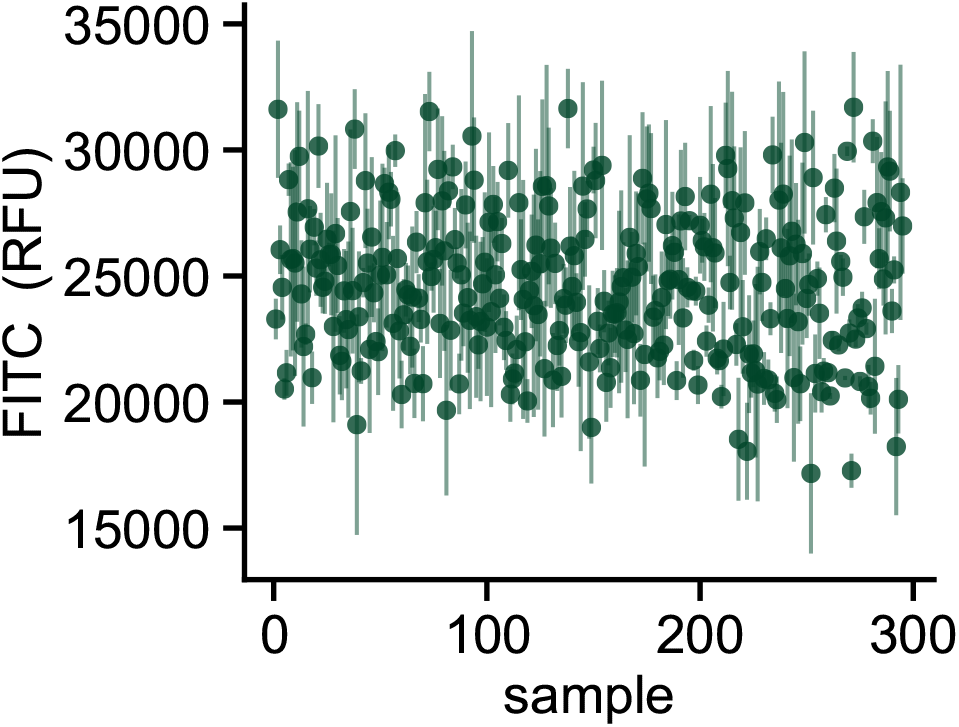
FITC spotting tracer. FITC-dextran (10 kDa) signal for each serum sample (1:8 dilution). Images were acquired in the spotting chamber after the sample had been resolubilized.

## Notes

### Competing Interest Statement

The authors have declared no competing interest.

### Author Declarations

Ethical approval was waived by the ethical committee of the University Hospital of Geneva in accordance with local regulations.

## References

[1] Zhou, P. et al. A pneumonia outbreak associated with a new coronavirus of probable bat origin. Nature 579, 270–273 (2020).

[2] Dong, E., Du, H. & Gardner, L. An interactive web-based dashboard to track COVID-19 in real time. The Lancet Infectious Diseases 20, 533–534 (2020).

[3] Perez-Saez, J. et al. Serology-informed estimates of SARS-CoV-2 infection fatality risk in Geneva, Switzerland. The Lancet Infectious Diseases (2020).

[4] Krammer, F. & Simon, V. Serology assays to manage COVID-19. Science 368, 1060–1061 (2020).

[5] Amanat, F. et al. A serological assay to detect SARS-CoV-2 seroconversion in humans. Nature Medicine 26, 1033–1036 (2020).

[6] Stringhini, S. et al. Seroprevalence of anti-SARS-CoV-2 IgG antibodies in Geneva, Switzerland (SEROCoV-POP): a population-based study. The Lancet 396, 313–319 (2020).

[7] Xu, X. et al. Seroprevalence of immunoglobulin M and G antibodies against SARS-CoV-2 in China.Nature Medicine 26, 1193–1195 (2020).

[8] Gudbjartsson, D. F. et al. Humoral Immune Response to SARS-CoV-2 in Iceland. New England Journal of Medicine (2020).

[9] Pollán, M. et al. Prevalence of SARS-CoV-2 in Spain (ENE-COVID): a nationwide, population-based seroepidemiological study. The Lancet 396, 535–544 (2020).

[10] Emmenegger, M. et al. Early peak and rapid decline of SARS-CoV-2 seroprevalence in a Swiss metropolitan region. medRxiv (2020).

[11] Norman, M. et al. Ultrasensitive high-resolution profiling of early seroconversion in patients with COVID-19. Nature Biomedical Engineering 1–8 (2020).

[12] Whitman, J. D. et al. Evaluation of SARS-CoV-2 serology assays reveals a range of test performance. Nature Biotechnology 1–10 (2020).

[13] Andrey, D. O. et al. Head-to-Head Accuracy Comparison of Three Commercial COVID-19 IgM/IgG Serology Rapid Tests. Journal of Clinical Medicine 9, 2369 (2020).

[14] Rudolf, F. et al. Clinical Characterisation of Eleven Lateral Flow Assays for Detection of COVID-19 Antibodies in a Population. medRxiv 2020.08.18.20177204 (2020).

[15] Maerkl, S. J. & Quake, S. R. A Systems Approach to Measuring the Binding Energy Landscapes of Transcription Factors. Science 315, 233–237 (2007).

[16] Garcia-Cordero, J. L. & Maerkl, S. J. Mechanically Induced Trapping of Molecular Interactions and Its Applications. Journal of Laboratory Automation 21, 356–367 (2016).

[17] Garcia-Cordero, J. L., Nembrini, C., Stano, A., Hubbell, J. A. & Maerkl, S. J. A high-throughput nanoimmunoassay chip applied to large-scale vaccine adjuvant screening. Integrative Biology 5, 650– 658 (2013).

[18] Garcia-Cordero, J. L. & Maerkl, S. J. A 1024-sample serum analyzer chip for cancer diagnostics. Lab on a Chip 14, 2642–2650 (2014).

[19] Thorsen, T., Maerkl, S. J. & Quake, S. R. Microfluidic Large-Scale Integration. Science 298, 580–584 (2002).

[20] Remy, M. M. et al. Effective chemical virus inactivation of patient serum compatible with accurate serodiagnosis of infections. Clinical Microbiology and Infection 25, 907.e7–907.e12 (2018).

[21] Fordyce, P. M. et al. De novo identification and biophysical characterization of transcription-factor binding sites with microfluidic affinity analysis. Nature Biotechnology 28, 970–975 (2010).

[22] Thevis, M. et al. Can dried blood spots (DBS) contribute to conducting comprehensive SARS-CoV-2 antibody tests? Drug Testing and Analysis 12, 994–997 (2020).

[23] Klumpp-Thomas, C. et al. Standardization of enzyme-linked immunosorbent assays for serosurveys of the SARS-CoV-2 pandemic using clinical and at-home blood sampling. medRxiv (2020).

[24] Garcia-Cordero, J. L. & Maerkl, S. J. Multiplexed surface micropatterning of proteins with a pressure- modulated microfluidic button-membrane. Chemical Communications 49, 1264–1266 (2013).

[25] Geertz, M., Shore, D. & Maerkl, S. J. Massively parallel measurements of molecular interaction kinetics on a microfluidic platform. Proceedings of the National Academy of Sciences 109, 16540– 16545 (2012).

[26] Piraino, F., Volpetti, F., Watson, C. & Maerkl, S. J. A Digital–Analog Microfluidic Platform for Patient- Centric Multiplexed Biomarker Diagnostics of Ultralow Volume Samples. ACS Nano 10, 1699–1710 (2016).

[27] Meyer, B. et al. Validation of a commercially available SARS-CoV-2 serological immunoassay. Clin- ical Microbiology and Infection 26, 1386–1394 (2020).

[28] Alvim, R. G. F. et al. An affordable anti-SARS-COV-2 spike protein ELISA test for early detection of IgG seroconversion suited for large-scale surveillance studies in low-income countries. medRxiv (2020).

[29] Chibwana, M. G. et al. High SARS-CoV-2 seroprevalence in Health Care Workers but relatively low numbers of deaths in urban Malawi. medRxiv (2020).

[30] Wrapp, D. et al. Cryo-EM structure of the 2019-nCoV spike in the prefusion conformation. Science 367, 1260–1263 (2020).

[31] Rockel, S., Geertz, M. & Maerkl, S. J. MITOMI: A Microfluidic Platform for In Vitro Characterization of Transcription Factor–DNA Interaction. Methods in Molecular Biology 786, 97–114 (2011).

[32] White, J. A. & Streets, A. M. Controller for microfluidic large-scale integration. HardwareX 3, 135– 145 (2017).

[33] Brower, K. et al. An Open-Source, Programmable Pneumatic Setup for Operation and Automated Control of Single and Multi-Layer Microfluidic Devices. HardwareX 1 – 37 (2017).

